# Peak alpha frequency as a neural marker of postoperative pain outcomes in spinal fusion surgery

**DOI:** 10.64898/2026.03.12.26348225

**Authors:** Amélie Grandjean, Fares Komboz, Tatjana Chacon, Lukas Weiser, Wolfgang Lehmann, Angelina Nazarenus, Dorothee Mielke, Veit Rohde, Ali Mazaheri, Tammam Abboud

**Affiliations:** School of Psychology, Centre for Human Brain Health, University of Birmingham, United Kingdom; Department of Neurosurgery, University Medical Center Göttingen, Germany; Department of Orthopedic Surgery, University Medical Center Göttingen, Germany; Department of Neurosurgery, University Medical Center Augsburg, Germany

**Author notes:** Tammam Abboud and Ali Mazaheri contributed equally to this work.

**Keywords:** Peak alpha frequency, electroencephalography, alpha oscillations, postoperative pain, spinal fusion surgery, pain prediction

## Abstract

Postoperative pain outcomes following spinal fusion surgery remain difficult to predict, as structural and surgical indicators alone offer limited insight into who will experience meaningful relief. A substantial proportion of patients continue to report persistent pain after surgery, underscoring the need for objective markers capable of identifying individuals at risk of poor recovery. Peak alpha frequency (PAF) has emerged as a promising trait-like neural signature of pain sensitivity. Experimental studies show that individuals with slower PAF exhibit heightened pain sensitivity. However, its ability to forecast longer-term postoperative pain trajectories remains unclear.

Seventeen adults undergoing cervical or lumbar fusion surgery were included. Resting-state, eyes-closed EEG was recorded preoperatively and at multiple visits after surgery. PAF was extracted from central electrodes using the centre-of-mass method. Pain intensity was assessed longitudinally using standardised self-report measures. Associations between PAF measures and postoperative pain change were examined using correlation analyses, and receiver operating characteristic (ROC) analyses evaluated discrimination of pain responders (≥50% improvement).

Preoperative PAF was positively associated with longer-term pain reduction at the 3-month follow-up, but showed no consistent relationship with early postoperative pain. Across pain measures, a consistent pattern emerged across the Brief Pain Inventory (BPI), Visual Analogue Scale (VAS), and Numerical Rating Scale (NRS), but not the Verbal Rating Scale (VRS) or Short-Form McGill (SF-MPQ). At three months, associations reached statistical significance for BPI-Worst (*ρ* = 0.67*, P* = 0.017), and BPI-Average Pain (*ρ* = 0.62*, P* = 0.033).

ROC analyses using BPI-Worst pain improvement demonstrated good discriminative ability of preoperative PAF for identifying treatment responders at 3 months (*AUC = 0.84; 95% CI: 0.61-1.00*), with high specificity and moderate sensitivity at the Youden-optimal threshold of 10.11 Hz. Changes in PAF over time were not reliably associated with changes in pain, suggesting that PAF functions primarily as a stable, trait-like predictor rather than a dynamic biomarker in this context.

This study demonstrates the feasibility and potential clinical value of preoperative EEG for characterising individual differences in postoperative pain recovery. The results identify faster preoperative PAF as a stable neural signal that captures meaningful variability in longer-term pain reduction, with convergent support across multiple patient-reported measures. While replication in a larger cohort is required, these findings establish a clear foundation for evaluating PAF as a candidate neurophysiological marker to inform preoperative risk profiling and potentially personalised perioperative pain-management strategies in spinal fusion patients.

## Introduction

Spine pain is one of the most prevalent musculoskeletal sources of pain globally^1^. Back pain is reported to affect 60-80% of all working adults at some point in their lives^2^ and is a leading cause of functional limitation and absence from work^3^. Both low back pain and neck pain are increasing in prevalence and combined are one of the largest contributors to duration of time living with a disability across countries and age ranges^4,5^. Beyond physical disability, chronic back and neck pain is also associated with poorer mental health^6^, highlighting the multifaceted impact of spine pain and the importance of understanding how to accurately diagnose and treat it.

### The clinical problem: mismatch between structural measures and symptoms

Spinal surgery is an established treatment for patients with neck or back pain caused by structural pathology such as stenosis, disc degeneration, or instability when conservative management fails^7–10^. Spinal fusion aims to stabilise affected segments and relieve neural compression to reduce pain^11–13^.

Surgical decision-making is typically based on a combination of symptoms, neurological findings, and imaging evidence of degenerative changes^5,8,14^. Despite substantial advances in imaging and surgical techniques^11,12^ structural correction does not reliably translate into pain relief. Degenerative changes are frequently observed in asymptomatic individuals, while symptomatic patients may show limited explanatory imaging findings^15,16^. This mismatch between structural measures and pain complicates patient selection and outcome prediction in spine surgery.

### Outcome variability: not all patients benefit from surgery

The discordance between structural pathology and symptoms extends to postoperative outcomes, with a substantial proportion of patients failing to achieve meaningful pain relief after spine surgery^17^. MRI-based measures of disease severity do not reliably predict pain or long-term improvement, surgical success is therefore primarily defined by patient-reported outcome measures rather than radiographic findings^18–20^.

Approximately 15-40% of patients experience insufficient improvement despite technically successful surgery^21^. While psychological and quality-of-life factors are associated with outcomes, they provide limited predictive accuracy^22,23^. Heterogeneous pain mechanisms and variable recovery trajectories further complicate outcome prediction and patient selection^15,24^.

### Importance of determining objective biomarkers

Imaging findings and psychological factors, while central to surgical planning, have not been shown to reliably predict postoperative pain outcomes following spinal fusion surgery^20,22^. Moreover, relying too heavily on psychological explanations can be poorly received by patients and may fail to reflect the biological mechanisms that contribute to pain persistence^25^. This highlights the need for objective biomarkers capable of distinguishing peripheral pathology from centrally mediated pain processes and improving individual pain-profiling approaches^26^.

A growing body of research has explored molecular, inflammatory, biomechanical, and integrative biomarkers of spine pain, including cytokines, inflammatory markers, immune phenotypes, and movement-based measures^27–31^. However, despite methodological advances and the use of data-driven approaches, no biomarker has yet demonstrated sufficient predictive accuracy for routine clinical use.

### Pain and the brain

Peripheral and biomechanical measures alone cannot explain the marked interindividual variability in pain experienced after comparable structural injury. Brain-based biomarkers offer direct insight into central pain modulation and the neural mechanisms that construct pain perception^32,33^. Cortical oscillations in the alpha frequency range (7-14 Hz) measured using EEG and magnetoencephalography (MEG) are closely linked to sensory gating and cortical excitability, processes consistently implicated in pain processing and vulnerability^34–38^. This has positioned EEG-derived measures as promising candidates for objective pain biomarkers, although clinical translation remains limited^39^.

The peak alpha frequency (PAF) is the dominant frequency within the alpha band. PAF is a trait-like measure with high test-retest reliability that reflects fundamental properties of brain function^40,41^. Slower PAF has been observed across several chronic pain populations and is associated with increased pain sensitivity and longer pain duration^42–45^. Importantly, baseline PAF measured before pain exposure predicts prolonged musculoskeletal pain, suggesting that PAF may reflect a pre-existing vulnerability rather than a consequence of pain^46,47^.

Clinically, a slower PAF may indicate that pain processing is more centralised rather than peripherally driven, and therefore less likely to improve through structural surgical intervention. Supporting feasibility, preoperative PAF has been shown to predict acute postoperative pain following thoracic surgery; a context where the outcome of interest is pain emerging after the procedure^48^. In contrast, spinal fusion surgery aims to reduce pre-existing pain through structural correction, and it is unknown whether PAF can predict pain improvement in this setting.

### Aims and hypotheses

This study investigates PAF as a biomarker of pain outcomes in patients undergoing cervical or lumbar fusion surgery. Specifically, we examined PAF as (i) a predictive biomarker, by testing whether preoperative PAF is associated with postoperative pain improvement, and (ii) a dynamic biomarker, by assessing whether longitudinal changes in PAF across postoperative visits are related to changes in pain.

Based on evidence that PAF reflects a stable, trait-like index of pain sensitivity and pain-relevant neural dynamics, we tested the following hypotheses:

1. Predictive hypothesis: Slower preoperative PAF is associated with smaller postoperative reductions in pain.
2. Dynamic hypothesis: Increases in PAF over postoperative follow-up visits are associated with greater improvements in pain, reflecting recovery of pain-relevant brain dynamics.

## Materials and methods

### Study design and population

This is a prospective, observational clinical study that was approved by the local Ethics Committee of Universitätsmedizin Göttingen (approval number: 11/6/23) and registered in the German Clinical Trials Register (DRKS00032164).

Only patients with a planned instrumented spinal fusion procedure for a degenerative disease of the cervical or lumbar spine were eligible for inclusion. Patient recruitment was conducted at the Departments of Neurosurgery and Orthopaedics, University Medical Center Göttingen.

Inclusion criteria were: (i) indication for surgical treatment of a degenerative spine disease requiring an instrumented fusion procedure; (ii) age between 18 and 80 years, and (iii) ability to understand the nature, significance, and scope of the study and to provide written informed consent.

Exclusion criteria included: severe internal medical or infectious disease, severe psychiatric illness, and current abuse of drugs, alcohol, or medication.

### Surgical procedure

Included patients underwent instrumented fusion of the cervical or lumbar spine, with or without additional decompression. The indication for spinal instrumentation was based on the presence of micro- or macroinstability of the affected spinal segment, as determined by clinical and radiological assessment. Patients treated with isolated decompression procedures or functional pain interventions were not included in this study.

### Participant assessments

Participants were assessed at four different time points. Visit 1 data were collected 72 to 24 hours preoperatively, Visit 2 data were collected 20 to 24 hours postoperatively, Visit 3 at discharge, and Visit 4 at 3-month follow-up. All study procedures are presented in Fig. 1.

**Figure 1:**
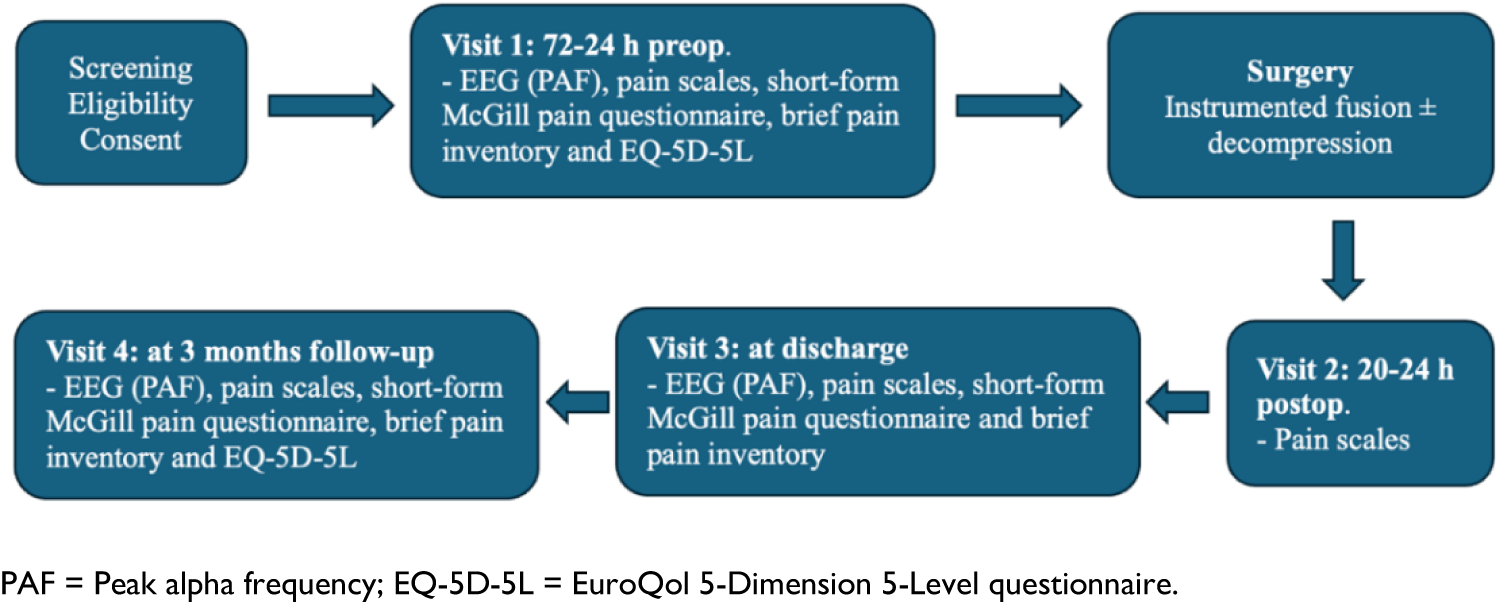
Flow diagram including the study visits and procedures.

### EEG recordings

Resting-state EEG was recorded at three time points: Preoperatively (72-24 hours before surgery), at hospital discharge, and at the 3-month follow-up visit.

Participants were seated and instructed to remain relaxed with their eyes closed throughout the recording. EEG data were acquired using ISIS Xpert® and NeuroExplorer 2019 (V. 5.1) software (Inomed Medizintechnik GmbH, Emmendingen, Germany) with eight scalp electrodes positioned according to the international 10-20 system. Signals were recorded at a sampling rate of 20,000 Hz with online filtering set to a high-pass filter 0.50 Hz, low-pass filter 70 Hz. Electrode impedances were maintained below 15 kΩ threshold for all channels. The duration of each resting-state recording was approximately 10 minutes.

### EEG preprocessing

The data were preprocessed using the EEGLAB toolbox^49^. Continuous recordings were first downsampled to 500 Hz, and the initial and final 3 seconds were removed to avoid edge artefacts. Line noise was removed using a 45-55 Hz notch filter, after which a 0.3 Hz high-pass and 40 Hz low-pass filter were applied. Each recording was then visually inspected to remove overly noisy segments of data due to movement. Across all sessions, the mean duration of rejected data per recording was 9.63 seconds (range: 0 - 72.55 seconds). A single channel was only removed from two recordings; otherwise, all present electrodes were retained. Finally, the datasets were re-referenced to the average and the online reference channel Cz was added back into the data.

### PAF

FieldTrip toolbox was used for frequency decomposition using the ft_freqanalysis function^50^ in MATLAB (version 2021a)^51^. The data were segmented into 5-second epochs, and power spectra computed for each individual epoch using a multitaper Fourier transform with a Hanning taper, yielding power estimates on a per-trial basis. Spectral estimates were calculated over a frequency range of 2-30 Hz.

PAF was estimated for each epoch using the centre-of-mass (CoM) method (also called the centre of gravity method) which calculates the weighted average frequency within the predefined alpha band. This approach has been used in previous pain research^48,52^ and is advantageous because it captures shifts in the distribution of alpha power rather than relying on a single spectral peak. Peak-picking can be unstable across epochs, as the maximum bin is susceptible to noise and may fluctuate substantially from trial to trial.

We defined as CoM as follows:

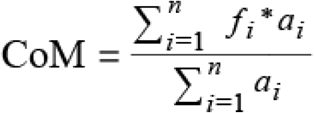

Where f_i_ is the ith frequency bin within the predetermined frequency range of interest of 7-14 Hz. n represents the 36 frequency bins ((14-7)/0.2 = 35, +1 for inclusion of end points), and a_i_ is the amplitude at the given f_i_. This is to say, each frequency within the 7-14 Hz range is multiplied by its power, the values are summed, then divided by the total power in the band, producing a weighted average that reflects the frequency at which alpha power is most concentrated.

PAF was calculated at the central region of interest comprising electrodes Cz, C3, and C4 due to alpha activity in sensorimotor regions predicting pain sensitivity in prolonged musculoskeletal pain^46,52^. Epoch-level central PAF values were then averaged over trials and central electrodes for a PAF estimate at each visit.

### Instruments for the assessment of pain

Although multiple validated pain measures were collected at each visit (outlined below), primary analyses focused on the Brief Pain Inventory (BPI)^53^. Unlike unidimensional intensity scales, the BPI provides a standardised, clinically meaningful assessment of both pain severity and its functional impact, which is particularly relevant in surgical populations where improvement is typically defined by reductions in pain and disability. Using the BPI as the primary outcome therefore offers a more comprehensive and interpretable measure of recovery while the additional scales support convergent validation of pain reports.

Pain assessments were conducted at all four study visits using established unidimensional pain scales:

#### Brief Pain Inventory (BPI)

The BPI is a validated questionnaire assessing pain severity and its impact on daily functioning. Analyses focused on the four pain severity items current pain, and worst, least, and average pain over the past 24 hours. The BPI is widely used in both clinical and research setting, and the worst pain item is of particular relevance to PAF-pain associations in surgical populations^48^.

#### Visual Analogue Scale (VAS)

Pain intensity was measured using a 100-mm VAS, a validated instruments widely used in clinical pain assessment^54,55^.

#### Numerical Rating Scale (NRS)

Participants also completed an 11-point NRS (0-10), a commonly used measure of pain, including in spine surgery research^56,57^. Although the NRS often corresponds with the VAS scores^58^, some research has shown the two scales are not interchangeable across all clinical populations^59^.

#### Verbal Rating Scale (VRS)

The VRS assesses pain using categorical descriptors (‘no pain’, ‘mild pain’, ‘moderate pain’, ‘serious/severe pain’, and ‘worst possible pain’). While some work shows the VAS and VRS may not be interchangeable in spine surgery populations, others demonstrate they are correlated in an orthopaedic clinical setting^60,61^.

#### Short-Form McGill Pain Questionnaire (SF-MPQ)

The SF-MPQ is a widely used multidimensional measure of pain experience with good reliability and validity across clinical pain populations^62^. It consists of 15 pain descriptors capturing sensory (11 items) and affective (4 items) dimensions of pain, each rated on a four-point intensity scale ranging from none to severe. Ratings can be summarized into sensory and affective subscale scores, as well as a total pain score reflecting overall pain quality.

### Quality of life

Health-related quality of life was assessed using the EQ-5D-5L questionnaire^63^ at the preoperative visit and at 3-month follow-up. Inclusion of quality of life measure aligns with IMMPACT recommendations, which emphasise outcomes in chronic pain research should extend beyond intensity and consider broader aspects of recovery^64^.

#### Hospital anxiety and depression

Anxiety and depression were measured using the Hospital Anxiety and Depression Scale (HADS), a 14-item questionnaire comprising separate anxiety and depression subscales (0–21 each)^65^.

#### Sample Size Justification

Sample size considerations were informed by Millard *et al*^48^ study reporting a strong association between preoperative PAF and postoperative pain intensity in a cohort of 16 thoracic surgery patients (*P* < 0.02)^48^. Although the exploratory nature and feasibility constraints of the present study limit the precision of a formal power calculation, the substantial effect size reported in Millard *et al*^48^ suggests that a sample of similar magnitude would be sufficient to detect an effect of comparable strength. We therefore set a minimum target enrolment of 16 patients, acknowledging that this sample size is intended to detect large effects and to provide preliminary evidence rather than definitive inferential conclusions.

### Statistical Analysis

#### Correlation analyses

Given evidence that 3-month patient-reported outcomes are the strongest and most consistent predictors of 12 month pain, disability, and satisfaction in spine surgery^66^, the change from preoperative assessment to 3-month follow-up is considered clinically most meaningful. Our primary correlation analyses examine change in pain (ΔPain) across this interval.

All statistical analyses were conducted in R (version 4.5.2)^67^. Because the patient-reported pain measures (BPI, VAS, and NRS) are widely considered to have ordinal measurement properties^58,68^, we prespecified Spearman’s rank correlations (*ρ*) as the primary tests for all PAF-pain associations. Pearson’s r was performed as a sensitivity analysis and is reported in the Supplementary Materials. The VRS provides categorical information only and was only assessed using Spearman’s rank correlations. This approach mitigates distributional assumptions and outlier sensitivity in our small sample while allowing comparison with prior clinical literature that sometimes treats pain scales as interval^69^.

We examined both directional change in PAF (ΔPAF) and absolute change in PAF (|ΔPAF|). Directional ΔPAF analyses were conducted with Spearman’s ρ (primary) and Pearson’s r (sensitivity), whereas magnitude (|ΔPAF|) was analysed analogously and flagged as exploratory. Full specifications are detailed in the Supplementary Materials.

#### Receiver Operating Characteristic (ROC) analyses

To examine the potential predictive value of PAF, ROC analyses were performed to determine the ability of preoperative PAF to distinguish treatment responders vs non-responders. Responders were defined based on change in pain outcomes between the preoperative visit and the 3-month follow-up. Analyses were performed using both a 30% improvement in pain and a 50% improvement as measured using the BPI-Worst, VAS, and NRS. These thresholds are based on IMMPACT consensus recommendations, clinically meaningful improvement in chronic pain conditions refers to ≥50% reduction in pain intensity as a “substantial” improvement, and a ≥30% as “moderately important” improvement^19^. Area under the curve (AUC) values with 95% confidence intervals were computed, with AUC ≥ 0.70 interpreted as fair discrimination and AUC ≥ 0.80 as good discrimination^70^.

#### Quality of life

EQ-5D-5L responses were converted into a single index value using the German valuation set^71^, following standard scoring procedures for deriving health-related quality-of-life utilities. EQ-VAS (EuroQol Visual Analogue Scale) were obtained as part of the EQ-5D questionnaire, representing self-rated health on a scale from 0 “worst imaginable health” to 100 “best imaginable health”. Changes in EQ-5D-5L and EQ-VAS from pre-op to follow-up were examined using non-parametric correlations with preoperative PAF.

## Results

### Patient characteristics

Between October 2023 and February 2025 seventeen adults undergoing spinal fusion were included in the study (mean age 59.5 ± 10.9, range: 37-75 years). The cohort comprised ten females (60.4 ± 10.1, range: 44-73 years) and seven males (58.3 ± 12.7, 37-75 years; Table 1). Fifteen participants underwent lumbar fusion, while two underwent cervical fusion. Of the cervical fusion cases, one participant was male and one was female.

**Table 1:** Mean ± standard deviations of baseline characteristics.

| Demographics | n | Mean (SD) | Min-Max |
| --- | --- | --- | --- |
| Age | 17 | 59.5 (10.9) | 37-75 |
| Sex | 17 | Male = 7 (41.2%) Female = 10 (58.8%) | - |

Five patients had previously undergone decompression of the affected spinal segments; no patient had a history of fusion surgery. Eleven patients underwent single-level fusion, whereas the remaining patients underwent multilevel fusion. All surgical procedures were uneventful, and postoperative CT imaging demonstrated correct implant positioning.

### Change in pain ratings from preoperative visit to three-month follow-up

Pain ratings showed a clear and consistent reduction from the preoperative visit to the 3-month follow-up across all measures. Scores on the BPI (Worst, Least, Average, and Current pain) decreased by approximately 63**-**71% (Table 2), and similar improvements were observed on both the VAS (-61%) and NRS (-63%, Table 3). By 3-month follow-up, several participants reported no pain, as reflected by minimum values of 0 across multiple scales. These reductions were also evident between patient discharge and three months, indicating continued improvement over the postoperative recovery period (Fig. 2). Overall, the data demonstrate a substantial decline in self-reported pain levels by three months after surgery.

**Figure 2:**
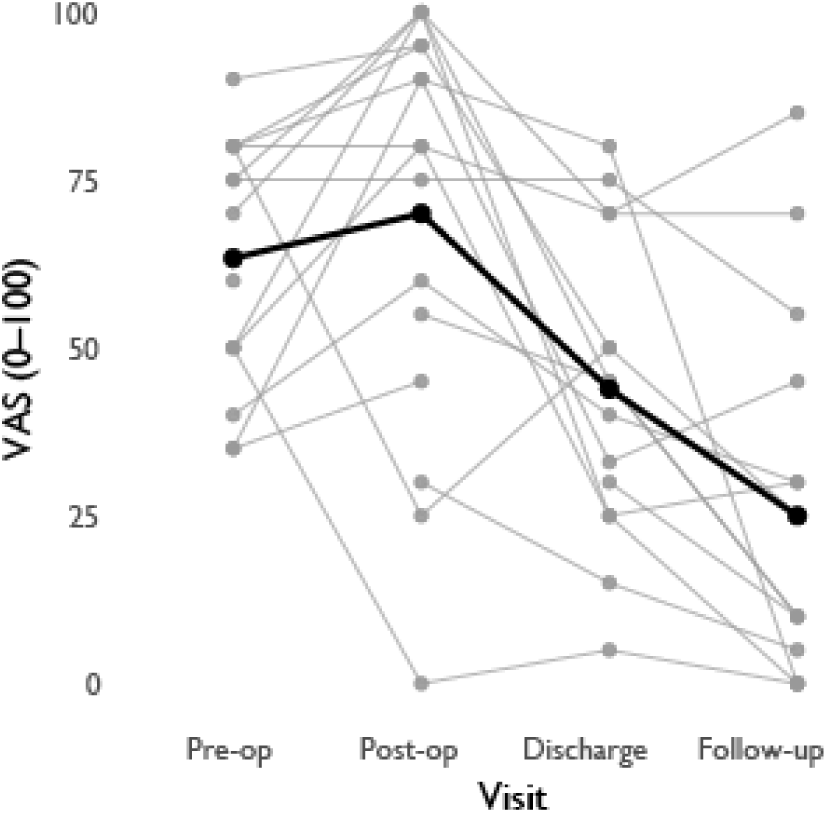
Pain trajectories across the 4 visits. VAS = Visual analogue scale; Pre-op = Visit 1 (72-24h before surgery); Post-op = Visit 2 (20-24h after surgery); Discharge = Visit 3; Follow-up = Visit 4 (three months after surgery). Grey lines represent individual participant pain scores, black line represents the group mean at each visit.

**Table 2:** Mean ± standard deviations on BPI at 3 time points.

|  | Pre-op |  | Discharge |  | Follow-up |  |
| --- | --- | --- | --- | --- | --- | --- |
| BPI Scale | n | Mean ± SD | n | Mean ± SD | n | Mean ± SD |
| **Worst** | 17 | 62.4 ± 31.9 | 15 | 43.3 ± 32.0 | 15 | 22.7 ± 31.0 |
| **Least** | 17 | 38.8 ± 32.2 | 15 | 30.7 ± 30.6 | 15 | 11.3 ± 17.7 |
| **Average** | 17 | 50.0 ± 28.1 | 15 | 38.7 ± 30.7 | 15 | 16.7 ± 23.2 |
| **Current** | 16 | 45.0 ± 30.6 | 15 | 36.0 ± 29.0 | 15 | 16.7 ± 24.7 |

BPI = Brief pain inventory; Pre-op = Visit 1 (72-24h before surgery); Discharge = Visit 3; Follow-up = Visit 4 (three months after surgery).

**Table 3:**
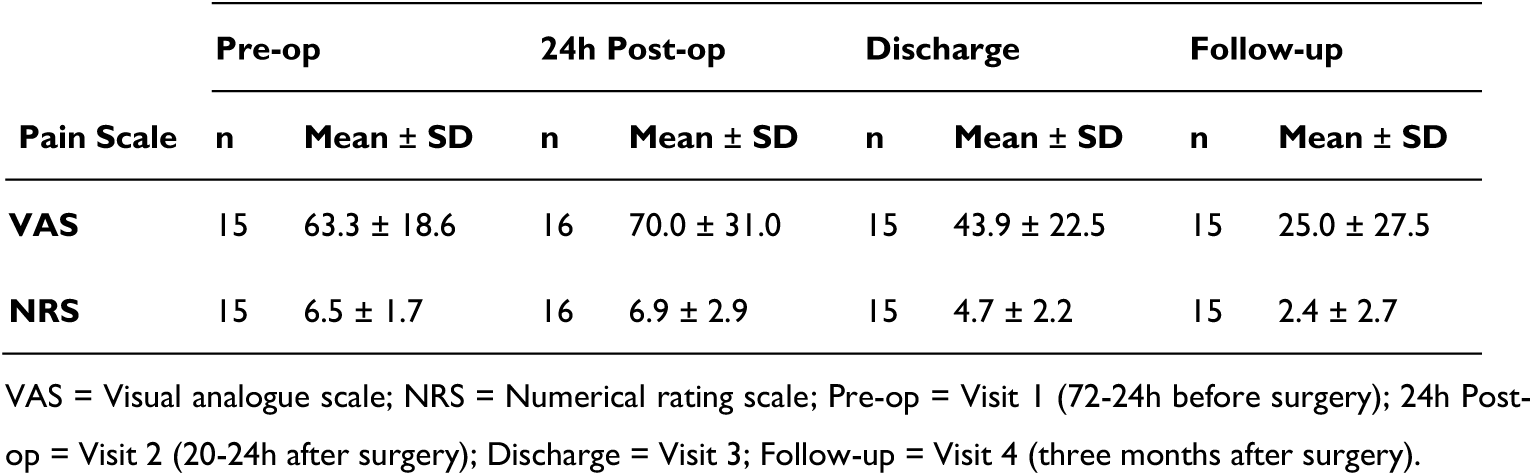
Mean ± standard deviations on VAS and NRS at 4 time points.

|  | Pre-op |  | 24h Post-op |  | Discharge |  | Follow-up |  |
| --- | --- | --- | --- | --- | --- | --- | --- | --- |
| Pain Scale | n | Mean ± SD | n | Mean ± SD | n | Mean ± SD | n | Mean ± SD |
| **VAS** | 15 | 63.3 ± 18.6 | 16 | 70.0 ± 31.0 | 15 | 43.9 ± 22.5 | 15 | 25.0 ± 27.5 |
| **NRS** | 15 | 6.5 ± 1.7 | 16 | 6.9 ± 2.9 | 15 | 4.7 ± 2.2 | 15 | 2.4 ± 2.7 |

VAS = Visual analogue scale; NRS = Numerical rating scale; Pre-op = Visit 1 (72-24h before surgery); 24h Post-op = Visit 2 (20-24h after surgery); Discharge = Visit 3; Follow-up = Visit 4 (three months after surgery).

On the VRS, most patients showed improvement by follow-up, with 8/15 improving, 5 unchanged, and none worsening (Table 4). For analysis and visualisation, VRS was coded numerically (0 = No pain, 1 = Mild, 2 = Moderate, 3 = Severe, 4 = Worst).

**Table 4:**
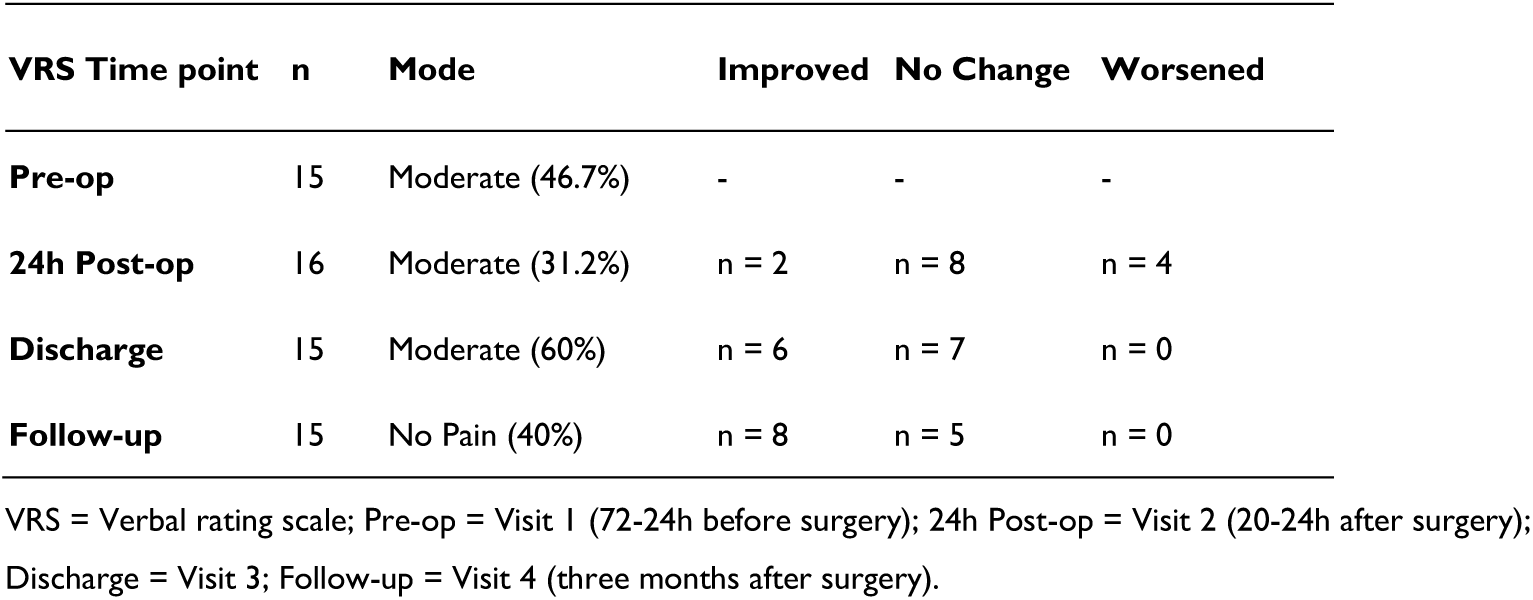
Mode and percentage improvement on VRS at 3 time points.

All pain scores were strongly intercorrelated at pre-op (*P* range = 0.62 - 0.98) indicating a substantial overlap across scales. At follow-up, intercorrelations were more heterogeneous (*P* range = 0.16 - 0.99). This reduction was primarily driven by weak associations involving the SF-MPQ affective-subscale. When this subscale was excluded, correlations at follow-up remained moderate to strong (*P* range = 0.52-0.99), with BPI-Least showing the weakest association with other scales (Supplementary Tables S4-S5). This pattern is not unexpected, given BPI-Least captures the minimum experience pain rather than typical or peak intensity. A larger separation between BPI-Least and other pain scales likely reflects greater variability in pain across time, whereas a smaller separation reflects more constant pain.

### Relationship between preoperative PAF and long-term change in pain ratings

By the 3-month follow-up, higher preoperative PAF was significantly associated with greater reductions in pain on BPI subscales (Fig. 3). Specifically, significant correlations were observed for BPI-Worst (*ρ* = 0.67, *P* = 0.017) and BPI-Average (*ρ* = 0.62, *P* = 0.033). BPI-Current showed a similar pattern, with a non-significant trend (*ρ* = 0.49, *P* = 0.122). BPI-Least demonstrated the weakest and non-significant trend (*ρ* = 0.28, *P* = 0.410). Sensitivity analyses using Pearson correlations supported the overall pattern, though not all effects reached significance, likely due to the modest sample size (Supplementary Table 12).

**Figure 3:**
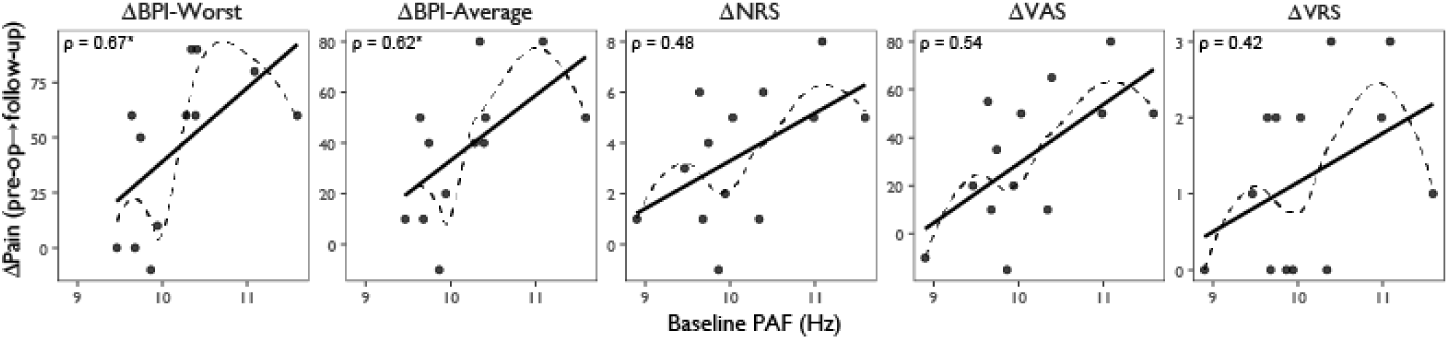
Associations between preoperative PAF and change in pain from preoperative visit to 3-month follow-up. Δ = Change (72-24h pre-operative visit minus three month follow-up visit); Baseline PAF = Peak alpha frequency at 72-24h preoperative visit; BPI-Worst = Brief pain inventory worst pain subscale BPI-Average = Brief pain inventory average pain subscale; NRS = Numerical rating scale; VAS = Visual analogue scale, VRS = Verbal rating scale. * Spearman rho correlation *P* <0.05.

Because patients reporting no pain prior to surgery could not show measurable improvement, they were excluded from the ΔBPI analyses. Three participants met this criterion, each reporting zero pain at baseline and at all subsequent visits, indicating a stable absence of pain rather than nonresponse. Analyses including these participants are provided in Supplementary Table S13.

For the VAS, NRS, and VRS, higher preoperative PAF tended to track with greater pain reduction at 3-month follow-up across all three scales. Associations were strongest for VAS (*ρ* = 0.54, *P* = 0.055) and were consistent but below the threshold for significance for NRS (*ρ* = 0.48, *P* = 0.096). Baseline PAF was not associated with change in VRS score (*ρ* = 0.42, *P* = 0.153).

Preoperative PAF did not predict change on the SF-MPQ. Across total, sensory, and affective scores, correlations for both pre-op to discharge and pre-op to follow-up were small and non-significant (*ρ* = 0.190 to 0.386; *P* = 0.155 to 0.498). Descriptive trajectories (pre-op, discharge, and follow-up) and full correlation outputs are provided in Supplementary Table S9 (descriptives) and Supplementary Table S10 (correlations).

### Relationship between preoperative PAF and short-term change in pain ratings

In contrast, only BPI-Least showed a significant relationship from pre-op to discharge (*ρ* = 0.62, *P* = 0.041). All correlations for BPI-Worst, BPI-Average, and BPI-Current were small and nonsignificant during this interval, indicating that PAF was not related to short-term changes in pain (*ρ* = -0.12, *P* = 0.709; *ρ* = -0.11, *P* = 0.739; *ρ* = 0.02, *P* = 0.946, respectively). Correlations were also weak and non-significant across all measures; VAS (*ρ* = 0.17, *P* = 0.579), NRS (*ρ* = 0.07, *P* = 0.828), and VRS (*ρ* = 0.25, *P* = 0.415).

Exploratory correlations were conducted for BPI-Impairment and perceived analgesic relief. A significant negative association was observed between baseline PAF and change in BPI-Impairment from pre-op to discharge (*ρ* = -0.79, *P* = 0.006). The association was not present at follow-up (*ρ* = 0.16, *P* = 0.619), indicating that this relationship was transient and specific to the early recovery phase. For perceived analgesic relief, all participants were retained in the analysis, as a baseline value of zero reflects absence of pain relief rather than absence of pain, and therefore represents a meaningful clinical state from which improvement can occur. No relationship between preoperative PAF and change in analgesic relief was observed at discharge (*ρ* = -0.46, *P* = 0.114) or follow-up (*ρ* = 0.09, *P* = 0.773).

### Sensitivity and specificity of preoperative alpha in predicting outcome category: responder vs non-responder

Prior to conducting the ROC analyses, we examined group-averaged preoperative power spectra for participants classified as responders and non-responders on the BPI-Worst subscale (≥50% improvement from pre-op to follow-up). This subscale was selected because it showed the strongest association with ΔPain in earlier analyses. Visual inspection revealed a clear rightward shift of the alpha peak in responders, indicating a faster dominant alpha rhythm relative to non-responders (Fig. 4). Consistent with this, the mean preoperative PAF was higher in responders (10.436 Hz) than in non-responders (9.738 Hz), suggesting meaningful spectral separation between outcome groups. These qualitative observations provide context for the formal classification results reported below.

**Figure 4:**
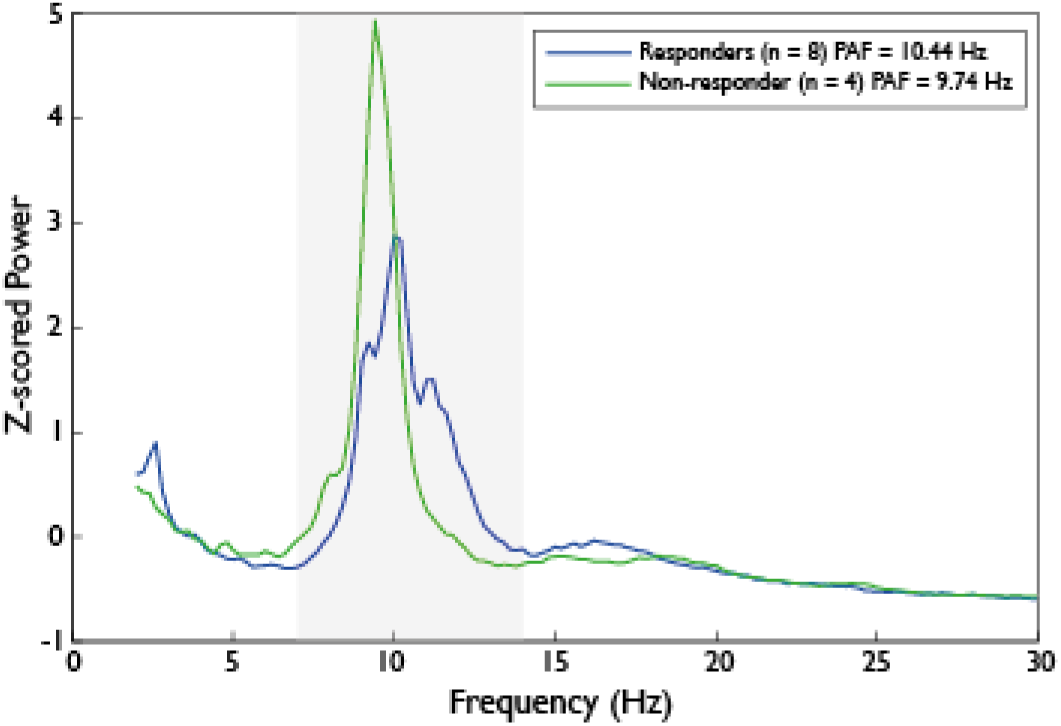
Preoperative Spectra: Responders vs Non-Responders. PAF = Peak alpha frequency; Responder = participants who showed ≥50% improvement in pain scores from pre-operative visit to 3-month follow up as measured using the BPI (Brief pain inventory). Non-responder = participants who showed <50% improvement in pain scores from pre-operative visit to 3-month follow up as measured using the BPI. Grey area represent the frequency range used to calculate PAF (7 – 14 Hz). Power spectra from central electrodes (Cz, C3, C4) were averaged for each participant and then converted to z-scores across frequencies to normalise differences in absolute spectral power. The resulting z-scored spectra were subsequently averaged across responders (n = 8) and non-responders (n = 4) for visualisation.

For ROC analyses, 12 patients had usable BPI-Worst data. Participants who reported zero baseline pain were excluded from percentage-based responder analysis to avoid misclassifying individuals with persistently zero pain as non-responders when no improvement was possible.

Using a ≥50% improvement threshold, preoperative PAF demonstrated good discrimination of responders on the BPI-Worst subscale (AUC = 0.84; 95% CI: 0.61, 1.00). The Youden optimal cut-off was 10.11 Hz, yielding a sensitivity of 0.75 and a specificity of 1.00 (Fig. 5).

**Figure 5:**
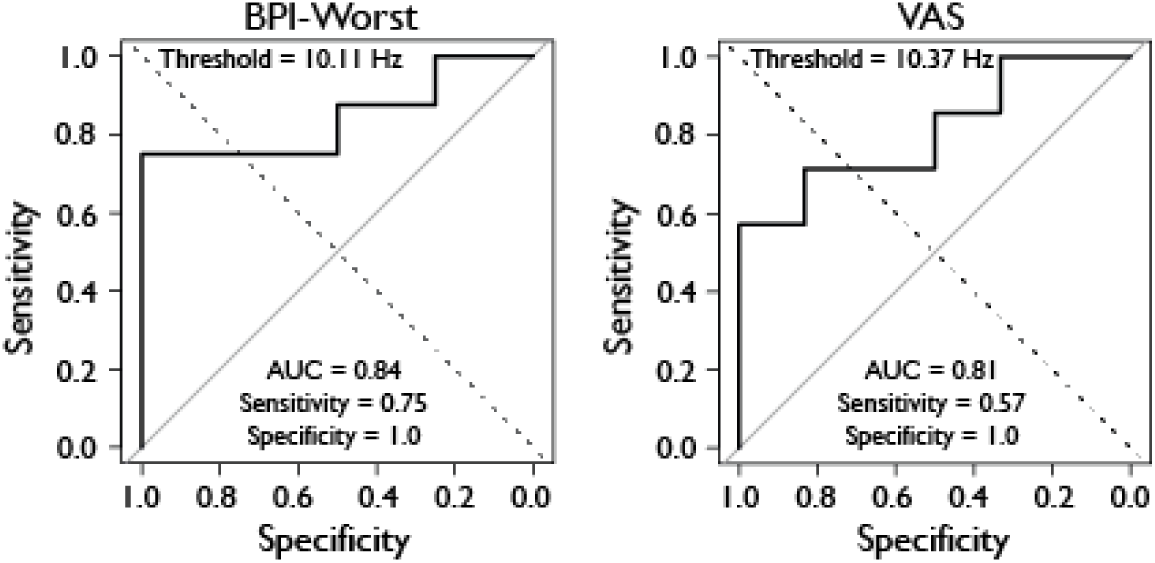
ROC curve for preoperative PAF predicting responders vs non-responders using the BPI-Worst and VAS. ROC curve = Receiver operating characteristic curve; PAF = Peak alpha frequency; BPI-Worst = Brief pain inventory worst pain subscale; VAS = Visual analogue scale; AUC = Area under the curve; Responder = participants who showed ≥50% improvement in pain scores from pre-operative visit to 3-month follow up as measured using the BPI (Brief pain inventory). Non-responder = participants who showed <50% improvement in pain scores from pre-operative visit to 3-month follow up as measured using the BPI or VAS.

Thirteen patients had complete data for VAS analyses. Preoperative PAF also showed good discriminative performance on the VAS (AUC = 0.81; 95% CI: 0.561, 1.00), again with high specificity (1.00) and moderate sensitivity (0.571). The optimal cut-off for VAS was 10.37 Hz. Notably, all individuals with preoperative PAF values above this threshold exhibited clinically meaningful improvement on both BPI-Worst and VAS, resulting in zero false-positive classifications.

Applying a more liberal ≥30% improvement threshold did not change responder classification for either BPI-Worst or VAS, and therefore produced identical ROC metrics. The NRS showed nearly identical results to the VAS; full NRS ROC outputs are presented in Supplementary Figs S4-S6.

To determine whether preoperative PAF discriminated responders from non-responders above chance (AUC = 0.5), we evaluated the statistical significance of each ROC curve. For the BPI-Worst scale, discrimination approached, but did not reach conventional significance (*P* = 0.07), despite a relatively high AUC. The VAS showed similar borderline evidence (*P =* 0.07-0.08), whereas no significant discrimination was observed for the NRS (*P* = 0.27). These results suggest potentially meaningful predictive value but also reflect limited power due to sample size and binary outcome classification. Full statistical procedures and supporting analyses are provided in the Supplementary Materials.

### Association between change in PAF and improvements in pain

ΔPAF from pre-op to discharge and pre-op to follow-up was not associated with ΔPain on the VAS, NRS, or BPI-Worst (all ρ’s non-significant). Cross-visit analyses (ΔPAF pre-op to discharge vs ΔPain pre-op to follow-up) were likewise null. Exploratorily, |ΔPAF| showed test-dependent patterns at pre-op to follow-up (Pearson significant for VAS/NRS and BPI-Average; Spearman non-significant), and should be interpreted cautiously. Full statistics are reported in Supplementary Tables S15-S16 (ΔPAF) and Supplementary Tables S17-S18 (|ΔPAF|), with scatterplots in Supplementary Fig. S7.

### Relationship between PAF and concurrent pain ratings

PAF did not correlate with concurrent pain at any visit (pre-op, discharge, follow-up) on VAS, NRS, VRS, or BPI subscales. Complete visit-wise correlation matrices are provided in Supplementary Table S19.

### Change in quality of life

Quality of life scores (EQ-5D-5L index and EQ-VAS) generally improved from pre-op to follow-up. Preoperative PAF showed no association with change in EQ-5D-5L index and a non-significant trend with change in EQ-VAS. Participant-level trajectories and statistics are in Supplementary Table S20.

### Hospital Anxiety and Depression Questionnaire

HADS-Anxiety and HADS-Depression decreased from pre-op to follow-up. Preoperative PAF showed a trend level association with change in anxiety (non-significant) and no association with change in depression. Full results appear in Supplementary Table S21.

### Opioid medication

Opioid medication was not analysed for outcome associations; individual PAF values and qualitative medication changes per visit are summarised in Supplementary Table S22.

## Discussion

This prospective observational study examined whether PAF from brief, pre-operative resting-state EEG can serve as a predictive (trait-like) biomarker of longer-term pain improvement after cervical or lumbar spinal fusion, and whether changes in PAF over time track changes in pain (dynamic marker). We observed that patients with faster baseline PAF were more likely to show meaningful pain reduction by 3 months, a pattern that was consistent across standard pain instruments. In contrast, PAF did not relate to early postoperative pain nor to multidimensional pain quality measures, and within-person changes in PAF did not reliably mirror changes in pain over time. Together, these findings position PAF as a stable, pre-surgical signal that helps characterise longer-term pain improvement rather than a dynamic biomarker for short-term pain or complex pain qualities.

### Long-term pain reduction is preferentially predicted by preoperative PAF

In support of the primary hypothesis, preoperative PAF was correlated with a reduction in pain at the 3-month follow-up. Change in pain was calculated by subtracting follow-up pain scores from preoperative pain scores; higher positive ΔPain scores indicated greater pain reduction. Faster preoperative PAF was related to greater reductions in worst and average pain, particularly on the BPI Worst, aligning with previous work showing that PAF relates most strongly to peak pain intensity^48^. ROC analyses support the predictive value of preoperative PAF. Using a responder definition based on ≥50% reduction in pain, faster baseline PAF demonstrated high specificity for identifying responders, indicating that individuals with a faster preoperative PAF were unlikely to be misclassified as responders when they did not in fact improve. Taken together, these findings suggest that PAF may serve as a probabilistic neurophysiological marker; individuals with faster PAF are more likely to show clinically meaningful pain improvement, though PAF alone does not deterministically predict postoperative outcomes.

We found that preoperative PAF did not relate to short-term postoperative pain changes (pre-op to discharge) on any measure with the exception of BPI-Least, indicating that baseline PAF is not strongly linked to early pain trajectories. Pain scores at discharge reflect the early recovery period, in which acute surgical injury, analgesic use, and inflammatory processes may dominate pain intensity, possibly influencing change in peak and average pain. In contrast, BPI-Least may be capturing the capacity to achieve periods of relative pain relief, which could be more sensitive to PAF as a trait-like difference in cortical inhibitory processes. As the acute postoperative recovery period subsides, these trait neurophysiological differences become broader across pain measures, as seen by the associations with change in pain at follow-up across various pain measures including peak pain captured by BPI-Worst.

Faster baseline PAF was related to smaller improvements on BPI-Impairment from the preoperative visit to discharge. This association was evident both when participants with zero baseline BPI-impairment were retained and when they were excluded, with larger effect sizes observed following exclusion. The association was not present at 3-month follow-up, indicating that this relationship was transient and specific to the early recovery phase. The direction of this effect contrasts with the positive association observed between baseline PAF and changes in pain intensity measures, suggesting that PAF may relate differently to early functional interference than to pain perception itself. One possible explanation is that individuals with faster baseline PAF may resume activity earlier in the postoperative period, resulting in greater perceived functional interference despite reductions in pain. Alternatively, this finding may reflect differing psychological or behavioural components captured by the BPI-impairment scale.

### PAF is not a real-time indicator of pain intensity

We found that PAF did not correlate with pain scores at any of the individual visits, on any of the scales. This suggests that PAF does not track how much pain a person is in at a given moment, and should not be interpreted as a real-time measure of how much pain someone is in. This is in line with findings that slower PAF in chronic pain does not indicate the intensity of pain, but may relate to other factors such as the degree of widespread pain^72^. Given that clinical observations suggest pre-operative pain intensity is a poor predictor of surgical outcomes^22^, a biomarker that predicts treatment response is not expected to correlate strongly with baseline subjective pain scores.

### Weak and exploratory associations between PAF shifts and pain outcomes

We did not find evidence that ΔPAF tracked ΔPain scores across visits. However, when examining absolute change from pre-op to follow-up, we found a weak association between |ΔPAF| and ΔPain using Pearson’s correlations. Although exploratory and dependent on the statistical test, this raises the possibility that individuals whose PAF shifts are more malleable over time may show greater improvements in pain.

PAF variability relating to change in pain is conceptually compatible with work suggesting different alpha generators may relate to pain sensitivity and coping strategies^73^. Furman *et al*^73^ propose that while the slower alpha generator may relate to heightened pain sensitivity^74^, the faster oscillator relates to a better tolerance for pain^45^ and that attenuation of the faster generator relates to efficient pain management. It is plausible that a “flexible” PAF may reflect a form of neural adaptability that supports better treatment response.

ΔPAF and |ΔPAF| from pre-op to discharge did not correlate with ΔPain from pre-op to follow-up, suggesting that the change in PAF from preoperative to postoperative cannot yet be considered a reliable marker for tracking postoperative recovery over a longer-term timescale.

### Individual variability in pain trajectories

In the current study, we observed substantial variability in postoperative pain trajectories. Pain scores generally trended down from preoperative to postoperative visits, although not all participants followed the same pattern. These findings were expected given that not all patients benefit from spine surgery^21^. Most participants showed some improvement, but the amount of change differed depending on the pain scale. On the VAS, two participants had higher pain scores at 3-month follow-up compared to pre-op, whereas on the NRS only one of these showed an increase. These findings therefore support the view that the VAS and NRS are not perfectly interchangeable and support the use of multiple tools to assess pain^59^. On the VRS, nobody showed an increase, but six of fifteen showed no change. Only one person reported an increase in pain on the BPI-Worst. The inter-individual variability in pain reduction trajectories directly motivates the investigation of why comparable surgical profiles can result in divergent pain outcomes.

Individual differences in PAF and PAF trajectories throughout the visits were also observed. Some participants displayed a reduction in PAF at the time of discharge, but their PAF returned to its baseline frequency at 3 months. The change at discharge may reflect increased postoperative pain. Although some evidence suggests PAF increases in acute experimental pain^74^, the present findings align with work demonstrating PAF slows in response to newly introduced sustained pain, which may better characterise the postoperative period^73^. PAF returning to a similar baseline for almost all participants supports the view that PAF represents a stable neurophysiological characteristic^40^.

### Possible mechanisms underlying the PAF-pain relationship

Several mechanisms may account for why higher preoperative PAF relates to postoperative pain improvement. First, the associations were observed at central electrodes, consistent with the role of alpha oscillations in regulating inhibition within sensorimotor networks. Alpha rhythms could suppress somatosensory and motor representations irrelevant to current goal-related networks^35^, and slower PAF has been linked to reduced inhibitory flexibility and heightened pain sensitivity^75^. From this perspective, individuals with slower PAF may have fewer neurophysiological resources to downregulate pain signals once tissue damage has been addressed, potentially hindering recovery.

A second interpretation centres on thalamocortical function. Slower PAF has been proposed to reflect disrupted thalamic relay or gating processes, which may contribute to amplification and maintenance of pain^76,77^. Individuals with faster baseline PAF may therefore exhibit more efficient thalamocortical processing, making them more likely to benefit from surgical correction of the peripheral pain source. Conversely, slower PAF could indicate greater central dysregulation, reducing the degree to which structural intervention translates into subjective pain relief.

This framework may also explain the exploratory observation that greater long-term modifiability of PAF relates weakly to improved pain outcomes. A more “flexible” PAF over the postoperative period may reflect thalamocortical circuits that adapt more effectively during recovery, aligning with accounts that different alpha generators relate to pain sensitivity versus coping capacities^73^. However, given the modest and statistically fragile effects, this interpretation remains tentative.

Finally, PAF may index broader cognitive-affective traits rather than pain-specific physiology. Previous work has linked PAF to anxiety, arousal, and attentional control^78–80^; factors known to influence postoperative trajectories^81^. Under this view, slower PAF may reflect a cognitive-affective profile that confers vulnerability to poorer recovery, whereas faster PAF may indicate greater resilience.

Collectively, these accounts suggest that PAF may capture a neurophysiological or cognitive profile that shapes an individual’s likelihood of benefiting from surgery, though the underlying mechanisms remain to be clarified.

### Summary and implications

This study provides preliminary evidence that PAF may capture individual differences in central pain processing that relate to postoperative improvement following spinal fusion surgery. In this cohort, faster preoperative PAF was probabilistically associated with greater long-term reductions in pain, supporting that PAF may index vulnerability to centrally maintained pain rather than reflecting pain intensity.

An important strength of PAF as a candidate biomarker is its clinical feasibility. We have demonstrated that PAF can be acquired rapidly and with minimal burden to patients, which makes it ideal for clinical translation. In addition, brain rhythms can be modulated. For example, neurofeedback, audio-visual stimulation, and non-invasive brain stimulation have been shown to influence oscillations, including alpha^82–84^. PAF may therefore not only represent a predictive marker, but also a target for future treatments aiming to improve postoperative outcomes.

### Limitations and future directions

The sample size was modest, and not all statistical associations reached significance. Future studies with larger sample sizes are necessary to confirm whether PAF reliably differentiates patients who will see meaningful improvements in pain scores from those who do not. One methodological consideration is that the VAS and NRS are sometimes considered ordinal, so a larger dataset would allow for continuous ordinal regression^68^. These modelling approaches would lead to a more rigorous assessment of the predictive value of PAF. In addition, preoperative pain may influence postoperative recovery^85^ and should be taken into consideration in future larger studies. PAF should not be viewed as an independent indicator, but is best thought of as a potential informative component within a multimodal framework that integrates clinical, psychological, and neurophysiological measures to improve prediction of treatment response^86^. Taken together, the present findings offer an encouraging proof-of-concept that PAF may serve as a meaningful neural marker of postoperative pain recovery after spinal fusion surgery.

## Data availability

The data generated and analysed during this study are available from the corresponding author upon reasonable request and with appropriate ethical approval.

## Acknowledgements

Ali Mazaheri is supported by funding from the Wellcome Leap Untangling Addiction Program.

