## Supplementary Materials for "Peak alpha frequency as a neural marker of postoperative pain outcomes in spinal fusion surgery"

#### Methods

##### PAF

EEG preprocessing procedures are described in detail in the main Methods; no additional preprocessing steps were applied for the supplementary analyses.

The frequency window of 7-14 Hz was selected to meaningfully capture all participants' alpha peaks at all time points. As shown in Fig. S1, overlaid central power spectra from all participants and all time points (pre-op, discharge, and 3-month follow-up) demonstrate inter-individual variability in peak frequency, spanning both slower and faster ranges. Importantly, alpha peak was observed for all participants. Narrow frequency windows would have failed to adequately encompass the full alpha peak for a subset of participants. The selected 7-14 Hz window consistently captured the complete alpha peak structure across individuals, rather than isolating only the local maximum. Inspection of the raw spectra further indicated no influence of aperiodic (1/f) activity, supporting the use of the wider window even when using the Centre of Mass method.

**Figure S1: Central power spectra across all participants and visits**

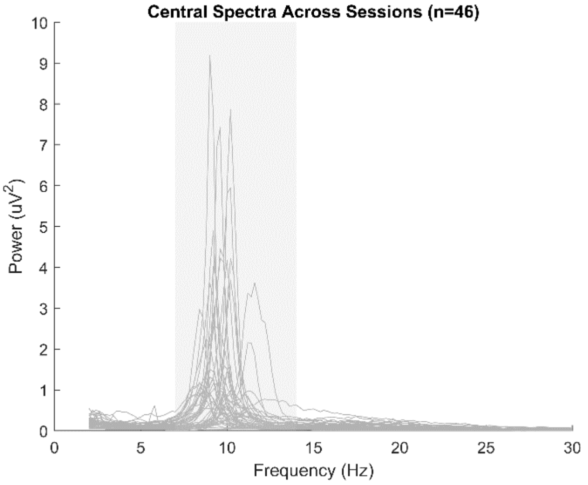


Each grey line represents the power spectrum of an individual participant at a single visit, averaged across central electrodes (Cz, C3, C4). Grey area represent the frequency range used to calculate PAF (7 – 14 Hz).

### Results

#### Descriptive statistics of alpha oscillations

Mean baseline PAF was 10.10 Hz (SD = 0.67, range 8.90–11.59). Although PAF values generally decreased at discharge (mean 9.92 Hz) and increased at follow-up (mean 10.02 Hz), individual variability in trajectories was observed (Fig. S2). For completeness, we also report alpha power across the time points, although no inferential analyses were conducted on alpha power as it does not form part of the primary hypotheses (Table S1). Alpha power reduced from pre-op (mean = 18.926) to discharge (mean = 15.753) and further decreased by 3-month follow-up (13.759).

**Table S1: Mean ± standard deviations of alpha oscillations at all time points**

| Alpha Measure | Visit | n | Mean ± SD |
|---|---|---|---|
| **PAF** | 1 | 17 | 10.099 ± 0.669 |
| | 3 | 16 | 9.915 ± 0.677 |
| | 4 | 13 | 10.023 ± 0.629 |
| **Alpha power** | 1 | 17 | 18.926 ± 15.455 |
| | 3 | 16 | 15.753 ± 9.315 |
| | 4 | 13 | 13.759 ± 10.647 |

PAF = peak alpha frequency (Hz); Alpha power = total power in the 7-14 Hz alpha band at central electrodes ($\mu V^2$)

For completeness, Supplementary Table S2 presents descriptive statistics of central PAF calculated using both the Centre of Mass (CoM) method and the Peak-picking approach. Peak-picking identifies the frequency within the alpha band (7-14 Hz) at which spectral power is maximal, whereas the CoM method computed a weighted average of the power across the alpha range, providing a more stable estimate. Consistent with the literature, more variability was observed using the Peak-picking method compared to CoM [1]. Estimates derived from CoM and Peak-picking methods were highly correlated at all study visits (Supplementary Table S3), indicating strong agreement between PAF calculation approaches. Given this high concordance and that CoM is more robust, all subsequent analyses focus on CoM derived PAF unless otherwise specified.

**Table S2: Mean ± standard deviations of alpha oscillations at all time points**

| Alpha Measure | Pre-op (n=17) | | Discharge (n=16) | | Follow-up (n=13) | |
|---|---|---|---|---|---|---|
| | Mean ± SD | Min-Max | Mean ± SD | Min-Max | Mean ± SD | Min-Max |
| **PAF - CoM** | 10.099 ± 0.669 | 8.901-11.591 | 9.915 ± 0.677 | 8.887-11.599 | 10.023 ± 0.629 | 8.864-11.236 |
| **PAF - Peak-picking** | 9.971 ± 0.963 | 8.527-12.015 | 9.702 ± 0.951 | 8.316-12.401 | 9.799 ± 0.830 | 8.360-11.604 |
| **Alpha Power** | 18.926 ± 15.455 | 1.415-50.483 | 15.753 ± 9.315 | 3.038-36.701 | 13.759 ± 10.647 | 2.061-37.098 |

PAF = Peak alpha frequency; CoM = Centre of Mass method of calculating PAF; Alpha power = total power in the 7-14 Hz alpha band at central electrodes ($\mu V^2$); Pre-op = Visit 1 (72-24h before surgery); Discharge = Visit 3; Follow-up = Visit 4 (three months after surgery).

**Table S3: Correlations between Centre of Mass and Peak-picking**

| Visit | n | Spearman ρ (p) | Pearson r (p) |
|---|---|---|---|
| **Pre-op** | 17 | 0.93 (5.6421e-08) | 0.97 (1.1841e-10) |
| **Discharge** | 16 | 0.94 (5.5889e-08) | 0.97 (5.1703e-10) |
| **Follow-up** | 13 | 0.93 (2.9987e-06) | 0.97 (2.372e-08) |

Pre-op = Visit 1 (72-24h before surgery); Discharge = Visit 3; Follow-up = Visit 4 (three months after surgery).

Mean PAF scores trended down from pre-op to discharge, then increased at follow-up. Individual variability was observed between participants, with 4 participants showing an increase from pre-op to discharge (Fig. S2). PAF values were normally distributed across the 3-visits (Fig. S3).

**Figure S2: Individual PAF trajectories across visits**

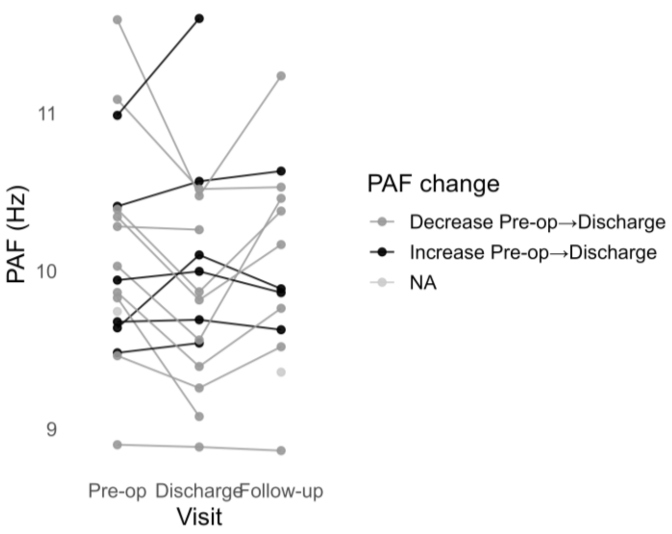


PAF = Peak alpha frequency; Pre-op = Visit 1 (72-24h before surgery); Discharge = Visit 3; Follow-up = Visit 4 (three months after surgery).

**Figure S3: PAF distributions at each visit**

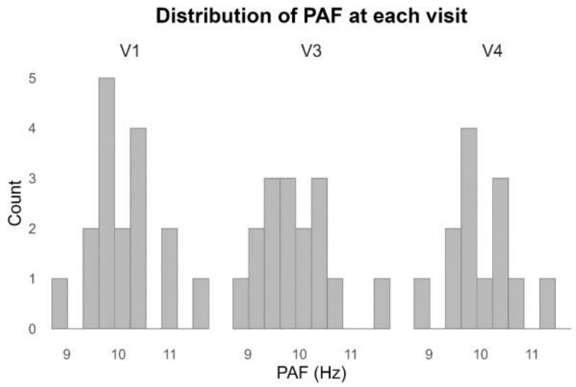


PAF = Peak alpha frequency; V1 = Visit 1 (72-24h before surgery); V3 = Visit 3 (discharge); V4 = Visit 4 (three months after surgery).

#### Pain scores were correlated with each other

All pain scores were strongly intercorrelated at pre-op (ρ range = 0.62 - 0.98) indicating a substantial overlap across scales (Table S4). At follow-up, intercorrelations were more heterogeneous (ρ range = 0.16 - 0.99; Table S5). This reduction was primarily driven by weak associations involving the SF-MPQ Affective subscale. When this subscale was excluded, correlations at follow-up remained moderate to strong (ρ range = 0.52-0.99), with BPI-Least showing the weakest association with other scales. This pattern is not unexpected, given that

BPI-Least captured minimum experienced pain, rather than typical or peak intensity. A larger separation between BPI-Least and other pain scales likely reflects greater variability in pain across time, whereas a smaller separation reflects more constant pain as might be expected before surgery compared to 3-month follow-up.

**Table S4: Pain scale intercorrelations at the pre-operative visit**

| | VAS | NRS | VRS | BPI-Worst | BPI-Average | BPI-Least | BPI-Current | SF-MPQ Total | SF-MPQ Affective | SF-MPQ Sensory |
|---|---|---|---|---|---|---|---|---|---|---|
| **VAS** | 1.00 | 0.96 | 0.91 | 0.80 | 0.81 | 0.87 | 0.84 | 0.72 | 0.78 | 0.71 |
| **NRS** | 0.96 | 1.00 | 0.92 | 0.75 | 0.82 | 0.86 | 0.84 | 0.72 | 0.73 | 0.72 |
| **VRS** | 0.91 | 0.92 | 1.00 | 0.81 | 0.81 | 0.88 | 0.89 | 0.62 | 0.69 | 0.63 |
| **BPI-Worst** | 0.80 | 0.75 | 0.81 | 1.00 | 0.78 | 0.74 | 0.75 | 0.81 | 0.83 | 0.81 |
| **BPI-Average** | 0.81 | 0.82 | 0.81 | 0.78 | 1.00 | 0.93 | 0.93 | 0.72 | 0.67 | 0.72 |
| **BPI-Least** | 0.87 | 0.86 | 0.88 | 0.74 | 0.93 | 1.00 | 0.98 | 0.73 | 0.73 | 0.69 |
| **BPI-Current** | 0.84 | 0.84 | 0.89 | 0.75 | 0.93 | 0.98 | 1.00 | 0.74 | 0.79 | 0.69 |
| **SF-MPQ Total** | 0.72 | 0.72 | 0.62 | 0.81 | 0.72 | 0.73 | 0.74 | 1.00 | 0.93 | 0.97 |
| **SF-MPQ Affective** | 0.78 | 0.73 | 0.69 | 0.83 | 0.67 | 0.73 | 0.79 | 0.93 | 1.00 | 0.85 |
| **SF-MPQ Sensory** | 0.71 | 0.72 | 0.63 | 0.81 | 0.72 | 0.69 | 0.69 | 0.97 | 0.85 | 1.00 |

VAS = Visual analogue scale; NRS = numerical rating scale; VRS = verbal rating scale; BPI = Brief pain inventory; SF-MPQ = Short-form McGill pain questionnaire.

**Table S5: Pain scale intercorrelations at 3-month follow-up**

| | VAS | NRS | VRS | BPI-Worst | BPI-Average | BPI-Least | BPI-Current | SF-MPQ Total | SF-MPQ Affective | SF-MPQ Sensory |
|---|---|---|---|---|---|---|---|---|---|---|
| **VAS** | 1.00 | 0.99 | 0.94 | 0.60 | 0.62 | 0.52 | 0.61 | 0.72 | 0.33 | 0.74 |
| **NRS** | 0.99 | 1.00 | 0.94 | 0.57 | 0.59 | 0.53 | 0.58 | 0.72 | 0.35 | 0.74 |
| **VRS** | 0.94 | 0.94 | 1.00 | 0.54 | 0.58 | 0.54 | 0.58 | 0.68 | 0.32 | 0.71 |
| **BPI-Worst** | 0.60 | 0.57 | 0.54 | 1.00 | 0.99 | 0.85 | 0.98 | 0.70 | 0.24 | 0.73 |
| **BPI-Average** | 0.62 | 0.59 | 0.58 | 0.99 | 1.00 | 0.87 | 0.99 | 0.70 | 0.21 | 0.73 |
| **BPI-Least** | 0.52 | 0.53 | 0.54 | 0.85 | 0.87 | 1.00 | 0.86 | 0.59 | 0.16 | 0.62 |
| **BPI-Current** | 0.61 | 0.58 | 0.58 | 0.98 | 0.99 | 0.86 | 1.00 | 0.70 | 0.23 | 0.73 |
| **SF-MPQ Total** | 0.72 | 0.72 | 0.68 | 0.70 | 0.70 | 0.59 | 0.70 | 1.00 | 0.75 | 0.98 |
| **SF-MPQ Affective** | 0.33 | 0.35 | 0.32 | 0.24 | 0.21 | 0.16 | 0.23 | 0.75 | 1.00 | 0.66 |
| **SF-MPQ Sensory** | 0.74 | 0.74 | 0.71 | 0.73 | 0.73 | 0.62 | 0.73 | 0.98 | 0.66 | 1.00 |

VAS = Visual analogue scale; NRS = numerical rating scale; VRS = verbal rating scale; BPI = Brief pain inventory; SF-MPQ = Short-form McGill pain questionnaire.

#### Change in pain ratings from preoperative visit to three-month follow-up

Across the BPI, VAS, NRS, and VRS, pain scores generally trended down from pre-op to follow-up. Supplementary Tables S6- S8 show the mean pain scores across the BPI, VAS, NRS, and VRS. One participant reported a pre-operative BPI-Least score that exceeded their reported BPI-Worst score, indicating a possible misunderstanding in their response. This participant did not contribute to longitudinal correlation analyses due to missing follow-up data; therefore, no sensitivity analyses were required.

**Table S6: Mean ± standard deviations on BPI at 4 time points**

| BPI Scale | n | Pre-op Mean ± SD | Pre-op Min-Max | n | Discharge Mean ± SD | Discharge Min-Max | n | Follow-up Mean ± SD | Follow-up Min-Max |
|---|---|---|---|---|---|---|---|---|---|
| Worst | 17 | 62.4 ± 31.9 | 0–90 | 15 | 43.3 ± 32.0 | 0–80 | 15 | 22.7 ± 31.0 | 0–80 |
| Least | 17 | 38.8 ± 32.2 | 0–100 | 15 | 30.7 ± 30.6 | 0–80 | 15 | 11.3 ± 17.7 | 0–60 |
| Average | 17 | 50.0 ± 28.1 | 0–90 | 15 | 38.7 ± 30.7 | 0–80 | 15 | 16.7 ± 23.2 | 0–70 |
| Current | 16 | 45.0 ± 30.6 | 0–90 | 15 | 36.0 ± 29.0 | 0–80 | 15 | 16.7 ± 24.7 | 0–70 |

BPI = Brief pain inventory; Pre-op = Visit 1 (72-24h before surgery); Discharge = Visit 3; Follow-up = Visit 4 (three months after surgery).

**Table S7: Mean ± standard deviations on VAS and NRS at 4 time points**

| | Pre-op (n = 15) | | 24h Post-op (n=16) | | Discharge (n=15) | | Follow-up (n=15) | |
|---|---|---|---|---|---|---|---|---|
| **Pain Scale** | **Mean ± SD** | **Min-Max** | **Mean ± SD** | **Min-Max** | **Mean ± SD** | **Min-Max** | **Mean ± SD** | **Min-Max** |
| **VAS** | 63.3 ± 18.6 | 35–90 | 70.0 ± 31.0 | 0-100 | 43.9 ± 22.5 | 5-80 | 25.0 ± 27.5 | 0-85 |
| **NRS** | 6.5 ± 1.7 | 4-9 | 6.9 ± 2.9 | 0-10 | 4.7 ± 2.2 | 2-8 | 2.4 ± 2.7 | 0-8 |

VAS = Visual analogue scale; NRS = Numerical rating scale; Pre-op = Visit 1 (72-24h before surgery); 24h Post-op = Visit 2 (20-24h after surgery); Discharge = Visit 3; Follow-up = Visit 4 (three months after surgery).

**Table S8: Mode and percentage of participants with improvement on VRS at 3 time points**

| Visit | n | Mode | Improved | No Change | Worsened |
|---|---|---|---|---|---|
| **Pre-op** | 15 | Moderate (46.7%) | - | - | - |
| **Post-op** | 16 | Moderate (31.2%) | n = 2 | n = 8 | n = 4 |
| **Discharge** | 15 | Moderate (60%) | n = 6 | n = 7 | n = 0 |
| **Follow-up** | 15 | No Pain (40%) | n = 8 | n = 5 | n = 0 |

VRS = Verbal rating scale; Pre-op = Visit 1 (72-24h before surgery); 24h Post-op = Visit 2 (20-24h after surgery); Discharge = Visit 3; Follow-up = Visit 4 (three months after surgery).

#### Relationship between PAF and McGill (SF-MPQ)

We also examined whether pre-operative PAF predicted change on the SF-MPQ. Although pain generally trended down from pre-op to follow-up (Table S9), no evidence of associations between PAF and SF-MPQ scores were found. Across all SF-MPQ total and subscale scores (n = 15), correlations at both pre-op to discharge and pre-op to follow-up were small and non-significant (Table S10).

**Table S9: McGill subscales at pre-op, discharge, and follow-up**

| SF-MPQ | Pre-op (n = 17) | | Discharge (n = 15) | | Follow-up (n = 15) | |
|---|---|---|---|---|---|---|
| **Subscale** | **Mean ± SD** | **Min-Max** | **Mean ± SD** | **Min-Max** | **Mean ± SD** | **Min-Max** |
| **Affective** | 4.412 ± 3.743 | 0 - 11 | 2.200 ± 2.111 | 0 - 7 | 1.800 ± 3.098 | 0 -11 |
| **Sensory** | 16.412 ± 8.132 | 4 - 28 | 9.733 ± 5.663 | 0 - 21 | 5.800 ± 6.816 | 0 - 22 |
| **Total** | 20.824 ± 11.408 | 5 - 37 | 11.933 ± 7.216 | 0 - 27 | 7.600 ± 9.440 | 0 - 29 |

SF-MPQ = Short-form McGill pain questionnaire; Pre-op = Visit 1 (72-24h before surgery); Discharge = Visit 3; Follow-up = Visit 4 (three months after surgery).

**Table S10: Correlations between preoperative PAF and change in McGill pain ratings**

| SF-MPQ | Pre-op→Discharge (n=15) | | Pre-op→Follow-up (n=15) | |
|---|---|---|---|---|
| **Subscale** | **Spearman ρ (p)** | **Pearson r (p)** | **Spearman ρ (p)** | **Pearson r (p)** |
| Affective | 0.270 (0.331) | 0.128 (0.649) | 0.380 (0.163) | 0.203 (0.469) |
| Sensory | 0.190 (0.498) | 0.183 (0.513) | 0.342 (0.212) | 0.319 (0.247) |
| Total | 0.247 (0.375) | 0.178 (0.525) | 0.386 (0.155) | 0.307 (0.266) |

SF-MPQ = Short-form McGill pain questionnaire; Pre-op = Visit 1 (72-24h before surgery); Discharge = Visit 3; Follow-up = Visit 4 (three months after surgery).

#### Relationship between PAF methods and change in all scales

For completeness, Table S11 reports the correlation values for baseline PAF and ΔPain on all scales, baseline PAF and ΔHADS, and baseline PAF and ΔEQ-5D-5L at V4.

**Table S11**: **Correlations between baseline PAF (CoM and Peak-picking) and change in pain, anxiety, depression, and quality of life from pre-op to follow-up**

| PAF Method | ΔPre-op →Follow-up Scale | n | Spearman ρ | Spearman p | Pearson r | Pearson p |
|---|---|---|---|---|---|---|
| **CoM** | **VAS** | 13 | 0.54 | 0.055 | 0.63 | *0.022** |
| **Peak-picking** | **VAS** | 13 | 0.46 | 0.114 | 0.61 | *0.026** |
| **CoM** | **NRS** | 13 | 0.48 | 0.096 | 0.53 | 0.06 |
| **Peak-picking** | **NRS** | 13 | 0.39 | 0.185 | 0.53 | 0.062 |
| **CoM** | **VRS** | 13 | 0.42 | 0.153 | 0.41 | 0.164 |
| **Peak-picking** | **VRS** | 13 | 0.31 | 0.302 | 0.4 | 0.176 |
| **CoM** | **BPI-Worst** | 12 | 0.67 | *0.017** | 0.58 | *0.049** |
| **Peak-picking** | **BPI-Worst** | 12 | 0.6 | *0.039** | 0.5 | 0.099 |
| **CoM** | **BPI-Least** | 11 | 0.28 | 0.41 | 0.39 | 0.23 |
| **Peak-picking** | **BPI-Least** | 11 | 0.38 | 0.252 | 0.45 | 0.162 |
| **CoM** | **BPI-Average** | 12 | 0.62 | *0.033** | 0.59 | *0.043** |
| **Peak-picking** | **BPI-Average** | 12 | 0.57 | 0.053 | 0.59 | *0.044** |
| **CoM** | **BPI-Current** | 11 | 0.49 | 0.122 | 0.63 | *0.039** |
| **Peak-picking** | **BPI-Current** | 11 | 0.5 | 0.118 | 0.61 | *0.044** |
| **CoM** | **SF-MPQ Total** | 15 | 0.39 | 0.155 | 0.31 | 0.266 |
| **Peak-picking** | **SF-MPQ Total** | 15 | 0.28 | 0.317 | 0.22 | 0.424 |
| **CoM** | **SF-MPQ Affective** | 15 | 0.38 | 0.163 | 0.2 | 0.469 |
| **Peak-picking** | **SF-MPQ Affective** | 15 | 0.23 | 0.408 | 0.1 | 0.734 |
| **CoM** | **SF-MPQ Sensory** | 15 | 0.34 | 0.212 | 0.32 | 0.247 |
| **Peak-picking** | **SF-MPQ Sensory** | 15 | 0.24 | 0.389 | 0.24 | 0.38 |
| **CoM** | **HADS Anxiety** | 15 | 0.5 | 0.056 | 0.47 | 0.074 |
| **Peak-picking** | **HADS Anxiety** | 15 | 0.42 | 0.115 | 0.49 | 0.067 |
| **CoM** | **HADS Depression** | 15 | 0.06 | 0.829 | 0.11 | 0.689 |
| **Peak-picking** | **HADS Depression** | 15 | 0.03 | 0.929 | 0.15 | 0.591 |
| **CoM** | **EQ-5D-5L** | 15 | 0.11 | 0.686 | 0.14 | 0.612 |

| | | | | | | |
|---|---|---|---|---|---|---|
| **Peak-picking** | **EQ-5D-5L** | 15 | -0.04 | 0.883 | 0.05 | 0.85 |
| **CoM** | **EQ-VAS** | 15 | 0.48 | 0.072 | 0.48 | 0.069 |
| **Peak-picking** | **EQ-VAS** | 15 | 0.36 | 0.187 | 0.39 | 0.151 |

PAF = Peak alpha frequency; CoM = Centre of Mass method of calculating; VAS = Visual analogue scale; NRS = numerical rating scale; VRS = verbal rating scale; BPI = Brief pain inventory; SF-MPQ = Short-form McGill pain questionnaire; HADS = Hospital anxiety and depression scale; EuroQol 5-Dimension 5-Level questionnaire; EQ-VAS = EuroQol Visual Analogue Scale.

#### Relationship between baseline PAF and change in BPI score

As reported in the main text, pre-operative PAF was significantly correlated with change in BPI-Worst and BPI-Average at the 3-month follow-up. Sensitivity analyses using Pearson's correlations confirmed these associations. For BPI-Current at 3 months, the association reached significance with Pearson's correlation but not with Spearman's. At discharge, the correlation between pre-operative PAF and change in BPI-Least was significant using Spearman's correlation; however, this association was not supported in the Pearson sensitivity analysis. A full summary of these results is presented in Table S12.

**Table S12. Correlations between preoperative PAF and change in BPI pain ratings**

| **BPI Scale** | **Pre-op →Discharge** | | | **Pre-op →Follow-up** | | |
|---|---|---|---|---|---|---|
| | **n** | **Spearman ρ (p)** | **Pearson r (p)** | **n** | **Spearman ρ (p)** | **Pearson r (p)** |
| **Worst** | 12 | -0.12 (0.709) | 0.02 (0.951) | 12 | *0.67 (0.017)** | *0.58 (0.049)** |
| **Least** | 11 | *0.62 (0.041)** | 0.50 (0.115) | 11 | 0.28 (0.410) | 0.39 (0.230) |
| **Average** | 12 | -0.11 (0.739) | 0.03 (0.915) | 12 | *0.62 (0.033)** | *0.59 (0.043)** |
| **Current** | 11 | 0.02 (0.946) | -0.02 (0.965) | 11 | 0.49 (0.122) | *0.63 (0.039)** |

PAF = Peak alpha frequency; BPI = Brief pain inventory. * $P \leq 0.05$.

To assess whether correlations between baseline PAF and changes in BPI outcomes were influenced by the inclusion or participants reporting no pain at both baseline and follow-up, analyses were repeated with these cases both retained and excluded. As shown in Table S13, the overall pattern of associations was largely consistent across both approaches. In particular, baseline PAF showed moderate positive correlations with change in BPI-Worsts and BPI-Average scores from pre-op to follow-up, with effect sizes generally increased when participants with zero baseline pain were excluded.

**Table S13**: **Correlations between baseline PAF and change in BPI-score from pre-op to follow-up**

| BPI-Scale | Keep baseline 0 y/n | n | Spearman ρ | Spearman p | Pearson r | Pearson p |
|---|---|---|---|---|---|---|
| **Worst** | y | 15 | *0.54* | *0.038** | 0.47 | 0.075 |
| **Worst** | n | 12 | *0.67* | *0.017** | *0.58* | *0.049** |
| **Least** | y | 15 | 0.39 | 0.147 | 0.40 | 0.144 |
| **Least** | n | 11 | 0.28 | 0.410 | 0.39 | 0.230 |
| **Average** | y | 15 | 0.46 | 0.085 | 0.48 | 0.073 |
| **Average** | n | 12 | 0.62 | *0.033** | *0.59* | *0.043** |
| **Current** | y | 14 | 0.45 | 0.105 | 0.44 | 0.113 |
| **Current** | n | 11 | 0.49 | 0.122 | 0.63 | 0.039 |

PAF = Peak alpha frequency; BPI = Brief pain inventory. * $P \leq 0.05$.

Exploratory correlations were conducted for BPI-Impairment and perceived analgesic relief. Results are reported here for completeness (Table S14). A significant negative association was observed between baseline PAF and change in BPI-Impairment from pre-op to discharge. This association was evident both when participants with zero baseline impairment were retained and when they were excluded, with larger effect sizes observed following exclusion. The association was not present at follow-up, indicating that this relationship was transient and specific to the early recovery phase.

For perceived analgesic relief, all participants were retained in the analysis, as a baseline value of zero reflects absence of pain relief rather than absence of pain, and therefore represents a meaningful clinical state from which improvement can occur. Results with both inclusion and exclusion of these participants are reported in Table S14. No significant associations were observed between baseline PAF and changes in perceived analgesic relief at either time window.

**Table S14**: **Exploratory analyses of the association between baseline PAF and change in BPI-Impairment and perceived analgesic relief**

| BPI-Scale | Keep baseline 0 y/n | Spearman ρ (p) | Pearson r (p) | Spearman ρ (p) | Pearson r (p) |
|---|---|---|---|---|---|
| **Impairment** | y | *-0.56 (0.038*)* | -0.52 (0.058) | 0.18 (0.523) | 0.23 (0.405) |
| **Impairment** | n | *-0.79 (0.006)** | -0.69 (0.018) | 0.16 (0.619) | 0.23 (0.465) |
| **Analgesic relief** | y | -0.46 (0.114) | -0.39 (0.184) | 0.09 (0.773) | 0.1 (0.762) |
| **Analgesic relief** | n | -0.11 (0.818) | -0.05 (0.915) | 0.6 (0.242) | 0.46 (0.355) |

PAF = Peak alpha frequency; BPI = Brief pain inventory. * *P* ≤ 0.05.

#### Sensitivity and specificity of pre-operative alpha in predicting outcome category: responder vs non-responder

When responders vs non-responders were defined using either the BPI-Worst or the VAS, baseline spectra showed comparable pattern, with non-responders exhibiting visibly slower PAF than responders (Fig. S4). The choice of outcome measure had minimal impact on group-level PAF estimates: using the VAS, mean baseline PAF was 9.699 Hz for non-responders and 10.496 Hz for responders, while using the BPI-Worst yielded similar values of 9.738 for non-responders and 10.436 Hz for responders.

**Figure S4: Pre-operative Spectra: Responders vs Non-Responders using BPI-Worst and VAS**

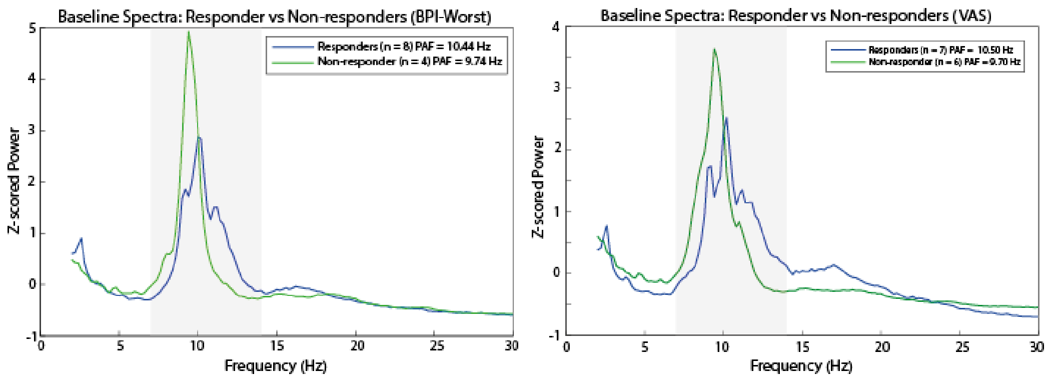


PAF = Peak alpha frequency; BPI-Worst = Brief pain inventory worst pain subscale; VAS = Visual analogye scale; Responder = participants who showed ≥50% improvement in pain scores from pre-operative visit to 3-month follow up as measured using the BPI and VAS. Non-responder = participants who showed <50% improvement in pain scores from pre-operative visit to 3-month follow up as measured using the BPI. Grey area represent the frequency range used to calculate PAF (7 – 14 Hz). Power spectra from central electrodes (Cz, C3, C4) were averaged for each participant and then converted to z-scores across frequencies to normalise differences in absolute spectral power. The resulting z-scored spectra were subsequently averaged across responders and non-responders for visualisation.

Using a 30% cut-off for BPI-Worst the threshold remained the same as the 50% threshold of 10.11 Hz. The AUC was 0.84 (95% CI: 0.607-1.00) with a sensitivity of 0.75 and specificity of 1. As with the 50% threshold, the calculated p value approached but did not reach statistical significance ($P$ = 0.073).

Thirteen patients had complete data for ROC curve analyses for the VAS and NRS. When applying the stricter 50% responder definition, the AUC for the VAS was 0.81 (95% CI: 0.561–1.00), and the NRS was 0.70 (95% CI: 0.394–1.00). The Youden-optimal cut-off was 10.37 Hz for both the VAS and NRS. Specificity remained high across both measures (1.00), with sensitivity ranging from 0.50 (NRS) to 0.571 (VAS) using the same 10.37 Hz cut-off. Using a 30% improvement threshold, pre-operative PAF showed fair discrimination for both VAS and NRS, with identical AUCs of 0.70 (95% CI: 0.394–1.00). The Youden-optimal cut-off was 10.37 Hz, yielding a specificity of 1.00 and a sensitivity of 0.50 for both measures. These near-identical ROC patterns reflect the underlying distribution of outcomes in this cohort: most participants either improved by more than 50% or by less than 30%, with few falling between these thresholds (VAS change: −28.6% to 100%; NRS change: −14.3% to 100%). Fig. S5 shows the ROC results for 50% improvement across BPI-Worst, VAS, and the NRS.

**Figure S5: ROC curve for preoperative PAF predicting responders vs non-responders using the BPI-Worst, VAS and NRS**

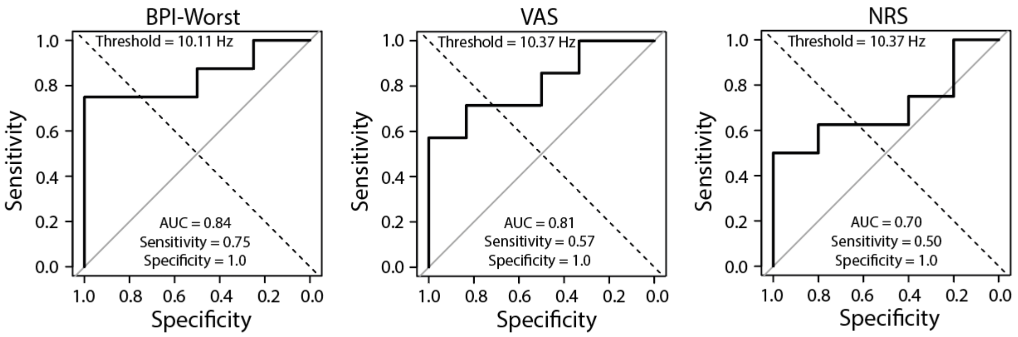


*ROC curve = Receiver operating characteristic curve; PAF = Peak alpha frequency; BPI-Worst = Brief pain inventory worst pain subscale; VAS = Visual analogue scale; AUC = Area under the curve;* Responder = participants who showed ≥50% improvement in pain scores from pre-operative visit to 3-month follow up as measured using the BPI (Brief pain inventory). Non-responder = participants who showed <50% improvement in pain scores from pre-operative visit to 3-month follow up as measured using the BPI, VAS, or NRS.

Overall, pre-operative PAF showed some potential to discriminate responder categories. Histograms of VAS, NRS, and BPI-Worst change scores showed no clear distributional structure. With the small sample size, the data were sparsely distributed across bins and did not exhibit normality (Fig. S6). Therefore, the predictive performance of PAF for responder category should be interpreted with caution.

**Figure S6**: **Histograms of change in pain from pre-op to follow-up on VAS, NRS, and BPI-Worst**

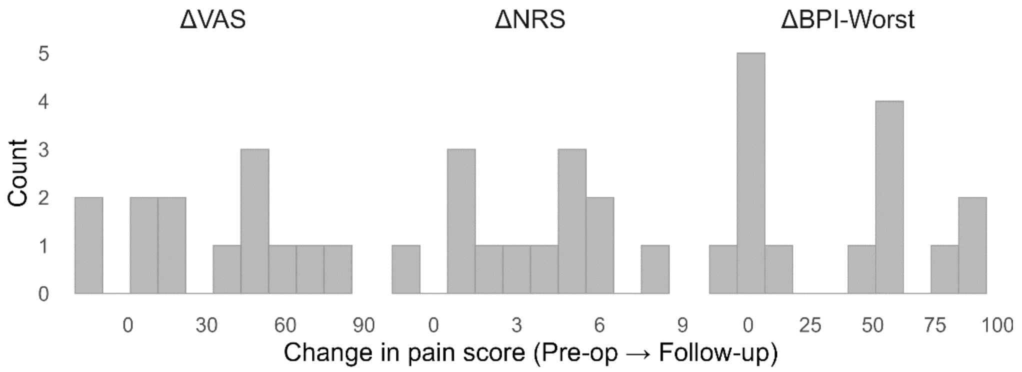


Δ = Change in pain (72-24h pre-operative visit minus three month follow-up visit); VAS = Visual analogue scale; NRS = Numerical rating scale; BPI-Worst = Brief pain inventory worst pain subscale. Pre-op = Visit 1 (72-24h before surgery); Follow-up = Visit 4 (three months after surgery).

To assess whether each AUC differed significantly from chance (AUC = 0.5), we used two mathematically equivalent non-parametric approaches: (i) a permutation test, in which responder labels were randomly permuted 10,000 times and an empirical p-value was computed as the proportion of shuffled datasets yielding an AUC ≥ the observed value; and (ii) a Wilcoxon rank-sum test, which compares PAF values between responders and non-responders and is analytically equivalent to testing whether AUC > 0.5 because the AUC represents the probability that a randomly selected responder has a higher PAF than a randomly selected non-responder. Both methods produced comparable results:

- BPI-Worst: permutation $P = 0.073$; Wilcoxon $P = 0.075$
- VAS: permutation $P = 0.08$; Wilcoxon $P = 0.07$
- NRS: permutation $P = 0.27$; Wilcoxon $P = 0.27$

These findings indicate that although AUCs were high (≥0.80 for BPI-Worst and VAS), statistical significance was not reached, reflecting limited power due to the small sample and dichotomisation of continuous outcomes.

#### Association between change in PAF and improvements in pain ratings

Change in PAF between visits was not associated with change in pain scores. For pre-op to discharge, correlations were weak and non-significant for VAS (ρ = 0.26, *P* = 0.38), NRS (ρ = 0.29, *P* = 0.339), and BPI-Worst (ρ = 0.34, *P* = 0.268). For pre-op to follow-up, there was no evidence of association between change in PAF and change in either VAS (ρ = 0.02, *P* = 0.948), NRS (ρ = 0.06, *P* = 0.853), or BPI-Worst (ρ = -0.07, *P* = 0.850). Using ΔPAF from pre-op to discharge tested against pain change from pre-op to follow-up also showed no evidence of association (VAS: ρ = −0.15, *P* = 0.65; NRS: ρ = −0.12, *P* = 0.72; BPI-Worst: ρ = -0.15, *P* = 0.661). No relationship between ΔPAF and ΔPain was observed on any of the BPI subscales. The cross-visit comparison was intended to test whether change in PAF by discharge could potentially be explored as a tool to predict final outcomes at 3-month follow-up. Across all comparisons, Pearson tests were consistent with Spearman, confirming no detectable relationship.

Tables S15 and S16 summarise correlations between changes in PAF and change in pain across early (pre-op to discharge) and later (pre-op to follow-up) postoperative intervals. Associations were examined both within the same time window and across windows, where early change in PAF was related to later changes in pain. For BPI outcomes, participants reporting zero pain at both baseline and follow-up were removed for these descriptive analyses.

**Table S15**: **Correlations between concurrent ΔPAF and ΔPain ratings**

| | Pre-op →Discharge | | | Pre-op →Follow-up | | |
|---|---|---|---|---|---|---|
| Pain Scale | n | Spearman ρ (p) | Pearson r (p) | n | Spearman ρ (p) | Pearson r (p) |
| **VAS** | 13 | 0.26 (0.385) | 0.21 (0.487) | 12 | 0.02 (0.948) | -0.12 (0.705) |
| **NRS** | 13 | 0.29 (0.339) | 0.16 (0.607) | 12 | 0.06 (0.853) | -0.11 (0.733) |
| **VRS** | 13 | 0.25 (0.415) | 0.28 (0.355) | 12 | 0.06 (0.865) | -0.06 (0.842) |
| **BPI-Worst** | 12 | 0.35 (0.268) | 0.21 (0.520) | 11 | -0.06 (0.851) | -0.14 (0.672) |
| **BPI-Least** | 11 | -0.19 (0.566) | -0.14 (0.692) | 10 | -0.22 (0.538) | -0.31 (0.387) |
| **BPI-Average** | 12 | 0.23 (0.479) | 0.06 (0.857) | 11 | -0.25 (0.451) | -0.34 (0.314) |
| **BPI-Current** | 11 | 0.17 (0.611) | 0.10 (0.769) | 10 | -0.24 (0.508) | -0.30 (0.403) |
| **SF-MPQ Sensory** | 15 | 0.22 (0.431) | 0.21 (0.458) | 13 | 0.11 (0.726) | 0.00 (0.994) |
| **SF-MPQ Affective** | 15 | 0.23 (0.413) | 0.21 (0.460) | 13 | 0.17 (0.584) | 0.19 (0.537) |

| | | | | | | |
|---|---|---|---|---|---|---|
| **SF-MPQ Total** | 15 | 0.31 (0.255) | 0.22 (0.431) | 13 | 0.15 (0.628) | 0.04 (0.895) |

ΔPain = Change in pain (72-24h pre-operative visit minus three month follow-up visit); ΔPAF = Change in PAF (three month follow-up visit minus 72-24h pre-operative visit); PAF = Peak alpha frequency; VAS = Visual analogue scale; NRS = numerical rating scale; VRS = verbal rating scale; BPI = Brief pain inventory; SF-MPQ = Short-form McGill pain questionnaire; Pre-op = Visit 1 (72-24h before surgery); Follow-up = Visit 4 (three months after surgery). Note: Positive ΔPAF values represent increase in PAF, while positive ΔPain values indicate reductions in pain. Higher positive correlations reflect greater pain improvements associated with larger increases in PAF.

**Table S16: Correlations between cross-visit ΔPAF (pre-op to discharge) and ΔPain (pre-op to follow-up)**

| Pain Variable | n | Spearman ρ (p) | Pearson r (p) |
|---|---|---|---|
| VAS | 12 | -0.14 (0.653) | -0.10 (0.768) |
| NRS | 12 | -0.12 (0.717) | -0.05 (0.879) |
| VRS | 12 | -0.15 (0.649) | -0.03 (0.932) |
| BPI-Worst | 11 | -0.15 (0.661) | -0.12 (0.718) |
| BPI-Least | 10 | -0.32 (0.370) | -0.18 (0.629) |
| BPI-Average | 11 | -0.29 (0.393) | -0.20 (0.548) |
| BPI-Current | 10 | -0.30 (0.400) | -0.15 (0.682) |
| SF-MPQ Sensory | 14 | -0.13 (0.663) | 0.01 (0.983) |
| SF-MPQ Affective | 14 | 0.28 (0.327) | 0.38 (0.179) |
| SF-MPQ Total | 14 | -0.12 (0.691) | 0.08 (0.777) |

ΔPain = Change in pain (72-24h pre-operative visit minus three month follow-up visit); ΔPAF = Change in PAF (discharge visit minus 72-24h pre-operative visit); PAF = Peak alpha frequency; VAS = Visual analogue scale; NRS = numerical rating scale; VRS = verbal rating scale; BPI = Brief pain inventory; SF-MPQ = Short-form McGill pain questionnaire; Pre-op = Visit 1 (72-24h before surgery); Discharge = Visit 3; Follow-up = Visit 4 (three months after surgery). Note: Positive ΔPAF values represent increase in PAF, while positive ΔPain values indicate reductions in pain. Higher positive correlations reflect greater pain improvements associated with larger increases in PAF.

Change in absolute PAF (|ΔPAF|) from pre-op to discharge was tested against pain improvement, defined as positive ΔBPI , ΔVAS, and ΔNRS (pre-op to discharge and pre-op to follow-up). At pre-op to discharge, no significant associations were observed (VAS: ρ = 0.41, *P* = 0.164; NRS: ρ = 0.24, *P* = 0.432; BPI-Worst: ρ = -0.31, *P* = 0.335).

For pre-op to follow-up, effect sizes increased (Fig. S7). For VAS, Spearman showed moderate but non-significant positive trends (ρ = 0.44, *P* = 0.153), while Pearson reached statistical significance (r = 0.63, *P* = 0.027). NRS results were consistent: Spearman ρ = 0.44 (*P* = 0.151), with Pearson significant at r = 0.64 (*P* = 0.024) (Table S17). For the BPI-subscales the BPI-

Average was significant with both Spearman's and Pearson's, (ρ = 0.63, *P* = 0.037, r = 0.65, *P* = 0.030) and BPI-Worst neared significance with Pearson's but not Spearman's (ρ = 0.45, *P* = 0.163, r = 0.565, *P* = 0.069).

**Figure S7: Associations between absolute change in PAF and change in pain from pre-op to follow-up**

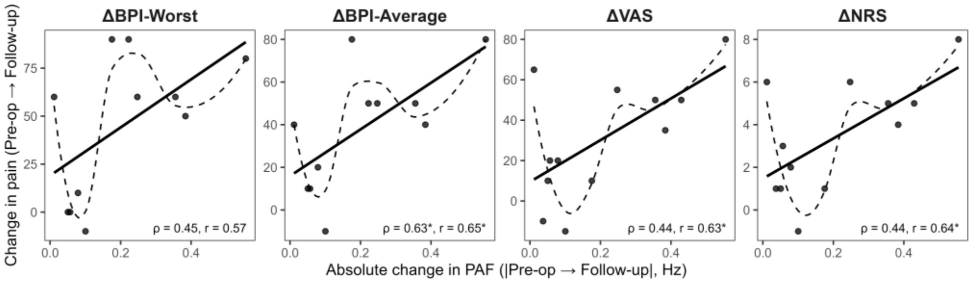


PAF = Peak alpha frequency; BPI-Worst = Brief pain inventory worst pain subscale; BPI-Average = Brief pain inventory average pain subscale; VAS = Visual analogue scale; NRS = Numerical rating scale; Pre-op = Visit 1 (72-24h before surgery); Follow-up = Visit 4 (three months after surgery). Note: change in pain calculated as 72-24h pre-operative visit minus three month follow-up visit.

Cross-visit analyses using |ΔPAF| from pre-op to discharge against pain change from pre-op to follow-up showed similar trends (VAS: ρ = 0.48, *P* = 0.117; r = 0.54, *P* = 0.069; NRS: ρ = 0.45, *P* = 0.141; r = 0.50, *P* = 0.102). This trend was not observed in the BPI-Worst (ρ = 0.374, *P* = 0.257; r = 0.35, *P* = 0.299) but neared significance on the other three subscales (Least: ρ = 0.59, *P* = 0.071; r = 0.35, *P* = 0.319; Average: ρ = 0.55, *P* = 0.074; r = 0.40, *P* = 0.224; Current: ρ = 0.60, *P* = 0.067; r = 0.55, *P* = 0.100).

Tables S17 and S18 report correlations between the magnitude of change in PAF (|ΔPAF|) and changes in pain outcomes across early and later intervals. Across measures, associations were generally modest and inconsistent, with some evidence of positive relationships between |ΔPAF| and ΔPain at follow-up.

**Table S17**: **Correlations between concurrent |ΔPAF| and ΔPain ratings**

| | Pre-op →Discharge | | | Pre-op →Follow-up | | |
|---|---|---|---|---|---|---|
| **Pain Scale** | **n** | **Spearman ρ (p)** | **Pearson r (p)** | **n** | **Spearman ρ (p)** | **Pearson r (p)** |
| **VAS** | 13 | 0.41 (0.164) | 0.43 (0.146) | 12 | *0.44 (0.153)* | *0.63 (0.027)** |
| **NRS** | 13 | 0.24 (0.432) | 0.30 (0.321) | 12 | *0.44 (0.151)* | *0.64 (0.024)** |
| **VRS** | 13 | 0.29 (0.339) | 0.18 (0.567) | 12 | 0.42 (0.176) | 0.56 (0.060) |
| **BPI-Worst** | 12 | -0.30 (0.355) | -0.13 (0.685) | 11 | 0.45 (0.163) | 0.57 (0.070) |
| **BPI-Least** | 11 | 0.52 (0.101) | 0.31 (0.356) | 10 | 0.36 (0.311) | 0.51 (0.133) |
| **BPI-Average** | 12 | -0.15 (0.640) | -0.01 (0.981) | 11 | *0.63 (0.037)** | *0.65 (0.030)** |
| **BPI-Current** | 11 | 0.16 (0.631) | 0.07 (0.848) | 10 | 0.52 (0.120) | 0.60 (0.066) |
| **SF-MPQ Sensory** | 15 | -0.11 (0.689) | -0.11 (0.690) | 13 | 0.38 (0.196) | 0.53 (0.064) |
| **SF-MPQ Affective** | 15 | 0.22 (0.425) | 0.00 (0.988) | 13 | 0.53 (0.063) | 0.47 (0.106) |
| **SF-MPQ Total** | 15 | -0.04 (0.893) | -0.08 (0.764) | 13 | 0.46 (0.117) | 0.53 (0.061) |

PAF = Peak alpha frequency; |ΔPAF| = absolute change in PAF from pre-op to follow-up; VAS = Visual analogue scale; NRS = numerical rating scale; VRS = verbal rating scale; BPI = Brief pain inventory; SF-MPQ = Short-form McGill pain questionnaire; Pre-op = Visit 1 (72-24h before surgery); Follow-up = Visit 4 (three months after surgery).

**Table S18**: **Correlations between cross-visit |ΔPAF| (pre-op to discharge) and ΔPain (pre-op to follow-up ratings)**

| **Pain Variable** | **n** | **Spearman ρ (p)** | **Pearson r (p)** |
|---|---|---|---|
| **VAS** | 12 | 0.48 (0.117) | 0.54 (0.069) |
| **NRS** | 12 | 0.45 (0.141) | 0.50 (0.102) |
| **VRS** | 12 | 0.50 (0.099) | 0.44 (0.153) |
| **BPI-Worst** | 11 | 0.37 (0.257) | 0.34 (0.299) |
| **BPI-Least** | 10 | 0.59 (0.071) | 0.35 (0.319) |
| **BPI-Average** | 11 | 0.56 (0.074) | 0.40 (0.224) |
| **BPI-Current** | 10 | 0.60 (0.067) | 0.55 (0.100) |
| **SF-MPQ Sensory** | 14 | 0.19 (0.526) | 0.20 (0.485) |
| **SF-MPQ Affective** | 14 | 0.09 (0.750) | 0.08 (0.779) |
| **SF-MPQ Total** | 14 | 0.19 (0.507) | 0.19 (0.052) |

PAF = Peak alpha frequency; |ΔPAF| = absolute change in PAF from pre-op to discharge; VAS = Visual analogue scale; NRS = numerical rating scale; VRS = verbal rating scale; BPI = Brief pain inventory; SF-MPQ = Short-form McGill pain questionnaire; Pre-op = Visit 1 (72-24h before surgery); Discharge = Visit 3; Follow-up = Visit 4 (three months after surgery).

#### Relationship between PAF and concurrent pain ratings

No evidence was found for an association between PAF and concurrent pain intensity at any visit over the VAS, NRS, VRS, or BPI. Preoperative PAF at pre-op was not related to pre-operative pain levels (VAS: ρ = 0.23, *P* = 0.406; NRS: ρ = 0.17, *P* = 0.551; VRS: ρ = 0.10, *P* = 0.734; BPI-Worst ρ = 0.17, *P* = 0.522). Similarly, PAF at discharge showed no relationship with pain ratings at discharge (VAS: ρ = -0.11, *P* = 0.703; NRS: ρ = -0.13, *P* = 0.638; VRS: ρ = -0.37, *P* = 0.180, BPI-Worst ρ = 0.06, *P* = 0.822), and follow-up PAF was not related to follow-up pain scores (VAS: ρ = -0.42, *P* = 0.149; NRS ρ = -0.41, *P* = 0.168; VRS: ρ = -0.39, *P* = 0.185, BPI-Worst ρ = -0.28, *P* = 0.363). Table S19 summarise associations. For BPI measures, all participants were retained in these analyses, as concurrent pain ratings were examined rather than change scores.

**Table S19**: **Concurrent PAF and pain scores at each visit**

| Pain Variable | Pre-op | | | Discharge | | | Follow-up | | |
|---|---|---|---|---|---|---|---|---|---|
| | n | Spearman ρ (p) | Pearson r (p) | n | Spearman ρ (p) | Pearson r (p) | n | Spearman ρ (p) | Pearson r (p) |
| **VAS** | 15 | 0.23 (0.406) | 0.23 (0.415) | 15 | -0.11 (0.703) | -0.26 (0.356) | 13 | -0.42 (0.149) | -0.40 (0.181) |
| **NRS** | 15 | 0.17 (0.551) | 0.12 (0.662) | 15 | -0.13 (0.638) | -0.22 (0.432) | 13 | -0.41 (0.168) | -0.33 (0.265) |
| **VRS** | 15 | 0.10 (0.734) | -0.03 (0.921) | 15 | -0.37 (0.180) | -0.42 (0.124) | 13 | -0.39 (0.185) | -0.36 (0.228) |
| **BPI-Worst** | 17 | 0.17 (0.522) | 0.14 (0.588) | 15 | 0.06 (0.822) | -0.03 (0.912) | 13 | -0.28 (0.363) | -0.29 (0.332) |
| **BPI-Least** | 17 | 0.23 (0.368) | 0.19 (0.459) | 15 | 0.03 (0.907) | -0.06 (0.827) | 13 | -0.06 (0.853) | -0.12 (0.700) |
| **BPI-Average** | 17 | 0.23 (0.372) | 0.18 (0.491) | 15 | 0.13 (0.638) | 0.03 (0.922) | 13 | -0.25 (0.401) | -0.25 (0.417) |
| **BPI-Current** | 16 | 0.16 (0.549) | 0.18 (0.512) | 15 | 0.19 (0.497) | 0.04 (0.878) | 13 | -0.23 (0.450) | -0.23 (0.445) |
| **SF-MPQ Total** | 17 | 0.08 (0.750) | 0.07 (0.785) | 15 | -0.26 (0.340) | -0.32 (0.240 | 13 | -0.03 (0.929) | -0.04 (0.890) |

PAF = Peak alpha frequency; VAS = Visual analogue scale; NRS = numerical rating scale; VRS = verbal rating scale; BPI = Brief pain inventory; SF-MPQ = Short-form McGill pain questionnaire; Pre-op = Visit 1 (72-24h before surgery); Discharge = Visit 3; Follow-up = Visit 4 (three months after surgery).

#### Change in quality of life

Health-related quality of life was assessed using the EQ-5D-5L questionnaire at each visit. The five dimensions (mobility, self-care, usual activities, pain/discomfort, and anxiety/depression) were coded according to the EuroQol Group's 5-level descriptive system (1 = no problems to 5 = extreme problems). Individual health states were converted into single index values using the German population-based value set [2] implemented through the official EQ-5D-5L algorithm. For each participant, change in EQ-5D-5L preference-based index score was calculated as ΔEQ-5D-5L = follow-up minus pre-op, such that positive values indicate improvement in overall health utility. Change in self-rated health was assessed using the EQ-5D-5L Visual Analogue Scale (EQ-VAS) calculated as ΔEQ-VAS = follow-up minus pre-op.

EQ-5D-5L index scores generally improved from pre-op to follow-up (mean change: 0.222, SD: 0.340) though five participants showed a decrease in index score over time (Table S20). EQ-VAS scores also demonstrated overall improvement (mean change: 21.667, SD: 21.715) with all but one participant reporting higher self-rated health at follow-up. This divergence suggests that changes in EQ-5D-5L preference-based index do not always align with participants' global self-assessment of their health, highlighting the EQ-VAS as a complementary measure capturing perceived overall health status on the day of assessment.

**Table S20: EQ-5D-5L at pre-op and follow-up, and change in EQ-5D-5L and EQ-VAS from pre-op to follow-up**

| | **EQ-5D-DL** | **EQ-5D_DL** | **Change in EQ-5D-DL** | **Change in EQ-VAS** |
|---|---|---|---|---|
| **ID** | **Pre-op** | **Follow-up** | **Follow-up – Pre-op** | **Follow-up – Pre-op** |
| **01** | 0.72 | 0.662 | -0.058 | -5 |
| **02** | 0.272 | 1 | 0.728 | 55 |
| **03** | 0.72 | 0.851 | 0.131 | 45 |
| **04** | 0.278 | 0.625 | 0.347 | 35 |
| **05** | 0.159 | 0.831 | 0.672 | 30 |
| **06** | 0.783 | 0.938 | 0.155 | 10 |
| **07** | 0.505 | - | - | - |
| **08** | -0.661 | - | - | - |
| **09** | 0.373 | 0.907 | 0.534 | 50 |
| **10** | 0.217 | 0.736 | 0.519 | 30 |
| **11** | 0.718 | 0.678 | -0.04 | 10 |

| | | | | |
|---|---|---|---|---|
| **12** | 0.298 | 0.816 | 0.518 | 45 |
| **13** | 0.242 | -0.22 | -0.462 | -10 |
| **14** | 0.441 | 0.375 | -0.066 | -10 |
| **15** | 0.298 | 0.502 | 0.204 | 20 |
| **16** | -0.003 | -0.165 | -0.162 | 7 |
| **17** | 0.543 | 0.856 | 0.313 | 13 |

EuroQol 5-Dimension 5-Level questionnaire; EQ-VAS = EuroQol Visual Analogue Scale; Pre-op = Visit 1 (72-24h before surgery); Follow-up = Visit 4 (three months after surgery).

In line with the observed difference between preference-based and self-rated health outcomes, baseline PAF was examined in relation to both EQ-5D-5L index score and EQ-VAS. Baseline PAF showed a moderate positive association with change in EQ-VAS from baseline to follow-up; however, this relationship did not reach statistical significance (Spearman $\rho = 0.48$, $P = 0.072$; Pearson $r = 0.48$, 95% CI −0.04 to 0.80, $P = 0.069$). In contrast, pre-operative PAF was not associated with change in EQ-5D-5L index scores. Correlations were small and non-significant across all tests (Spearman $\rho = 0.11$, $P = 0.69$; Pearson $r = 0.14$, $P = 0.613$). These findings suggest that PAF may preferentially relate to pain and perceptual aspects of recovery rather than broader multidimensional evaluations of overall wellbeing.

#### Hospital Anxiety and Depression Questionnaire

HADS scores showed an overall reduction from pre-op to follow-up. Mean anxiety scores decreased from 9.47 (SD = 3.62; range = 1-15) at pre-op to 7.76 (SD = 3.13; range = 0-17) at follow-up. Similarly, mean depression scores decreased from 6.20 (SD = 4.87; range = 3-12) at pre-op to 5.47 (SD = 3.76; range = 0-14) at follow-up.

Descriptive statistics for the HADS are provided in Table S21, showing mean, standard deviation, and range of anxiety and depression scores at pre-op and follow-up. These data are included to characterise the distribution and range of affective symptoms in the cohort across time.

**Table S21**: **HADS scores at pre-op and follow-up**

| | Pre-op (n = 17) | | Follow-up (n = 15) | |
|---|---|---|---|---|
| **HADS Scale** | **Mean ± SD** | **Min-Max** | **Mean ± SD** | **Min-Max** |
| **Anxiety** | 9.47 ± 3.62 | 1-15 | 7.76 ± 3.13 | 0-17 |
| **Depression** | 6.20 ± 4.87 | 3-12 | 5.47 ± 3.76 | 0-14 |

HADS = Hospital anxiety and depression scale; Pre-op = Visit 1 (72-24h before surgery); Follow-up = Visit 4 (three months after surgery).

Pre-operative PAF showed a trend-level association with change in anxiety symptoms, such that higher preoperative PAF showed a trend level association with greater reductions in HADS-Anxiety scores which did not reach statistical significance ($\rho = 0.5$, $P = 0.056$; $r = 0.47$, $P = 0.074$). No relationship was observed between pre-operative PAF and change in depressive symptoms on the HADS-Depression subscale ($\rho = 0.06$, $P = 0.829$, $r = 0.11$, $P = 0.689$).

#### Opioid medication

To document medication changes over time, Table S22 presents increases or decreases in opioid use. Opioid use was not analysed.

**Table S22: Individual opioid use at all time points**

| ID | Pre-op | 24h Post-op | ΔPre-op →24h Post-op | Discharge | Δ24h Post-op →Discharge | Follow-up | Δ24h Post-op →Follow-up |
|---|---|---|---|---|---|---|---|
| **01** | N | Y | Increase | Y | Decrease | N | Decrease |
| **02** | N | Y | Increase | N | Decrease | N | Decrease |
| **03** | N | Y | Increase | Y | Change * | N | Decrease |
| **04** | Y | Y | Increase | Y | Decrease | - | - |
| **05** | N | N | No change | N | No change | N | No change |
| **06** | Y | Y | - | N | Decrease | N | Decrease |
| **07** | N | Y | Increase | Y | No change | - | - |
| **08** | Y | Y | No change | Y | Increase | - | - |
| **09** | N | Y | Increase | Y | Decrease | N | Decrease |
| **10** | Y | Y | Increase | Y | Decrease | Y | Decrease |
| **11** | N | Y | Increase | Y | Increase | Y | Increase |
| **12** | N | Y | Increase | Y | Decrease | Y | Decrease |
| **13** | Y | Y | Increase | Y | Increase | N | Decrease |

| | | | | | | | |
|---|---|---|---|---|---|---|---|
| **14** | N | Y | Increase | Y | Increase | N | Decrease |
| **15** | N | Y | Increase | N | Decrease | N | Decrease |
| **16** | Y | N | Decrease | N | No change | N | No change |

Pre-op = Visit 1 (72-24h before surgery); Post-op = Visit 2 (20-24h after surgery); Discharge = Visit 3; Follow-up = Visit 4 (three months after surgery). *Change in opioid medication not possible to ascertain total increase or decrease as decreased Oxycodone but started Acetylsalicylsäure and Metamizol.
